# Ten Years of Scientific Discovery with the UK Biobank CMR Imaging Study

**DOI:** 10.64898/2025.12.20.25342751

**Authors:** Zahra Raisi-Estabragh, Steffen E. Petersen, Stefan Neubauer

**Author notes:** **Address for correspondence:** Zahra Raisi-Estabragh; William Harvey Research Institute, NIHR Barts Biomedical Research Centre, Queen Mary University of London, Charterhouse Square, London, EC1M 6BQ. **Funding:** SN acknowledges support from the Oxford National Institute for Health Research Biomedical Research Centre and the Oxford British Heart Foundation Centre of Research Excellence. SEP acknowledges the support of the National Institute for Health Research Barts Biomedical Research Centre (NIHR203330). **Disclosures:** SN is a Perspectum shareholder and is a joint holder of patents related to the use of MRI for the assessment of liver disease. SEP provides consultancy to Cardiovascular Imaging Inc, Calgary, Alberta, Canada. **Acknowledgements:** We would like to acknowledge Dr Andrew Tyler (University of Manchester, UK Biobank Ltd) for creating Figures 1 and 2, and Ms Jackie Cooper (Queen Mary University of London) for creating Figures 5 and 6. The magnetic resonance images in Figures 1 and 2 are used with permission of UK Biobank. The central illustration was created using Biorender.

## Abstract

The UK Biobank Imaging Study, with its dedicated cardiovascular magnetic resonance (CMR) sub-study, has re-defined the scale and scope of cardiovascular research, generating high-quality imaging data in 100,000 participants with linkage to rich genetic, demographic, lifestyle, and clinical data. The resource has enabled transformative discoveries across genomics, epidemiology, and biomedical engineering, and has served as a global blueprint for population imaging studies. Its success has been accelerated by an equitable data access model that fosters international collaboration. Looking ahead, efforts should focus on harmonisation across cohorts, adherence to rigorous methodological standards, and multidisciplinary collaboration to drive meaningful clinical translation – whether through direct improvements in patient care or experimental validation of imaging-derived insights. The UK Biobank CMR experience illustrates the power of large-scale imaging cohorts and sets a benchmark for future initiatives aimed at improving cardiovascular health through integrated, collaborative science. This paper provides an overview of the UK Biobank and its CMR sub-study, systematically reviews key publications, discusses methodological considerations, and highlights important future directions.

## Introduction

In the 20^th^ century, public health strategies drove significant advancements in population health and life expectancy^1^. The most striking achievements were in the prevention and control of infectious diseases. Life expectancy in developed nations increased more during this time than in any previous period. As population demographics shifted towards greater longevity and burden from infectious diseases declined, non-communicable conditions, particularly cardiovascular diseases (CVDs), emerged as the predominant public health challenge^2^. By the 1940s, CVD had become the leading cause of mortality among Americans, accounting for 1 in 2 deaths^3^.

The Framingham Heart Study was established in 1948 as the first longitudinal study of CVD, enrolling over 5,000 residents of the city of Framingham (United States)^3^. The study was the first to present robust evidence for major cardiovascular risk factors, such as smoking, high cholesterol, hypertension, and low physical activity^3^. Modification of these factors continues to shape the foundations of preventive cardiology today.

In subsequent years, large-scale longitudinal cohorts were established across the world, incorporating different populations and varying approaches to participant characterisation, such as the Million Veteran Program (Unites States)^4^ and the China Kadoorie Biobank (China)^5^. Improvements in electronic health data infrastructures permitted health record linkage for many cohorts, allowing capture of documented health outcomes from registry records and other health databases. These developments broadened the scope of possible studies and strengthened their methodological rigour and statistical power.

In parallel, there was increasing interest in identification of pre-clinical disease states, where preventive strategies or early therapeutic interventions may be implemented to improve prognostic outlook. The search for early CVD biomarkers remains a major research priority.

Cardiovascular imaging is at the core of cardiovascular clinical care, informing diagnostic and treatment decisions across the spectrum of cardiology. In the context of population research, cardiovascular imaging provides continuous measures of cardiac anatomy, function, and physiology – thereby presenting highly granular definitions of risk across the entire sample enhancing statistical power and enabling more precise mapping of disease trajectories, especially in the early and intermediate stages.

Cardiovascular magnetic resonance (CMR) is the reference modality for cardiac chamber quantification and provides unique non-invasive myocardial tissue characterisation^6,7^. Over the last two decades, rapid technical advancements in CMR have generated new understanding of many cardiovascular conditions. Early CMR studies were focused on feasibility in small cohorts of patients and volunteers^8,9^. As the technique became more established, larger studies were undertaken focused on detailed characterisation of distinct clinical cohorts^10,11^, with some expanding to create dedicated registries – such as the Hypertrophic Cardiomyopathy (HCM) Registry^12^. It was also increasingly recognised that remodelling patterns defined using CMR often preceded the onset of clinically manifest disease^13,14^, highlighting the potential of CMR biomarkers for enhanced CVD detection and prediction.

More recently, multimodal biomedical databases have emerged, which, for the first time, incorporate large-scale CMR imaging – such as, the Multi-Ethnic Study of Atherosclerosis (MESA, United States)^15^, the German National Cohort (Germany)^16^, and the UK Biobank (United Kingdom)^17^ – presenting new opportunities for cardiovascular research. Among these, the UK Biobank comprises the world’s largest CMR databank presented alongside detailed participant phenotyping and linkages to national health record databases.

This paper provides an overview of the UK Biobank and its CMR sub-study, a systematic review of its contributions to cardiovascular research, methodological considerations, and directions for future research.

### Background to UK Biobank

The UK Biobank is one of the largest longitudinal cohort studies. A total of 9.2 million invites were posted to individuals aged 40-69 years old living within 25 miles of one of 22 assessment centres across the United Kingdom, identified through National Health Service registers^18^. There were 503,317 respondents (5.5% response rate), with recruitment undertaken between 2006 to 2010^19^. All participants provided written informed consent. There was no pre-requisite for healthy status at baseline, but individuals who were unable to provide consent or complete baseline assessment due to ill-health or discomfort were not included.

Baseline assessment comprised a touchscreen questionnaire, verbal interview, a series of physical measures, and collection of biological samples (blood, saliva, urine). This resulted in extensive characterisation of socio-demographics, lifestyle factors, medical history, and biomarker profiling for all participants. Multiple aliquots of biological samples were stored for future analysis. Approximately 20,000 participants were recalled for repeat assessment in a calibration visit between 2012 to 2013.

Linkages are established with the Office for National Statistics (ONS) mortality records, Hospital Episode Statistics (HES), and the National Cancer Registration and Analysis Service (NCRAS) database, capturing health events recorded in accordance with standardised International Classification of Disease (ICD) codes. This information is periodically refreshed by the UK Biobank, permitting longitudinal tracking of incident health events for the entire cohort.

The UK Biobank Imaging Study started in 2014 with the aim of scanning 100,000 of the original participants^17,20^ – in 2025, this important landmark was achieved. All scans were performed in one of four dedicated imaging centres (Cheadle, Reading, Newcastle, and Bristol) using uniform and standardised equipment, staff training, acquisition parameters, and quality control procedures. The protocol includes multiorgan imaging with magnetic resonance scans of the heart, brain, and abdomen, carotid ultrasound, and whole-body DEXA (dual energy x-ray absorptiometry). A protocol for management of incidental findings was also developed, informed by evidence gathered during the pilot phase of the study^21^, and consultations with clinical, ethical, and legal stakeholders – with consensus to provide limited feedback for findings considered to be potentially likely to threaten life span, quality of life, or major body functions observed during data acquisition or quality control.

Repeat imaging for 60,000 participants began in 2019 and is scheduled for completion in 2029. Example images from the UK Biobank multimodal multi-organ imaging assessment are provided in **Figure 1**. The information collected at baseline assessment is repeated at each imaging visit.

**Figure 1.**
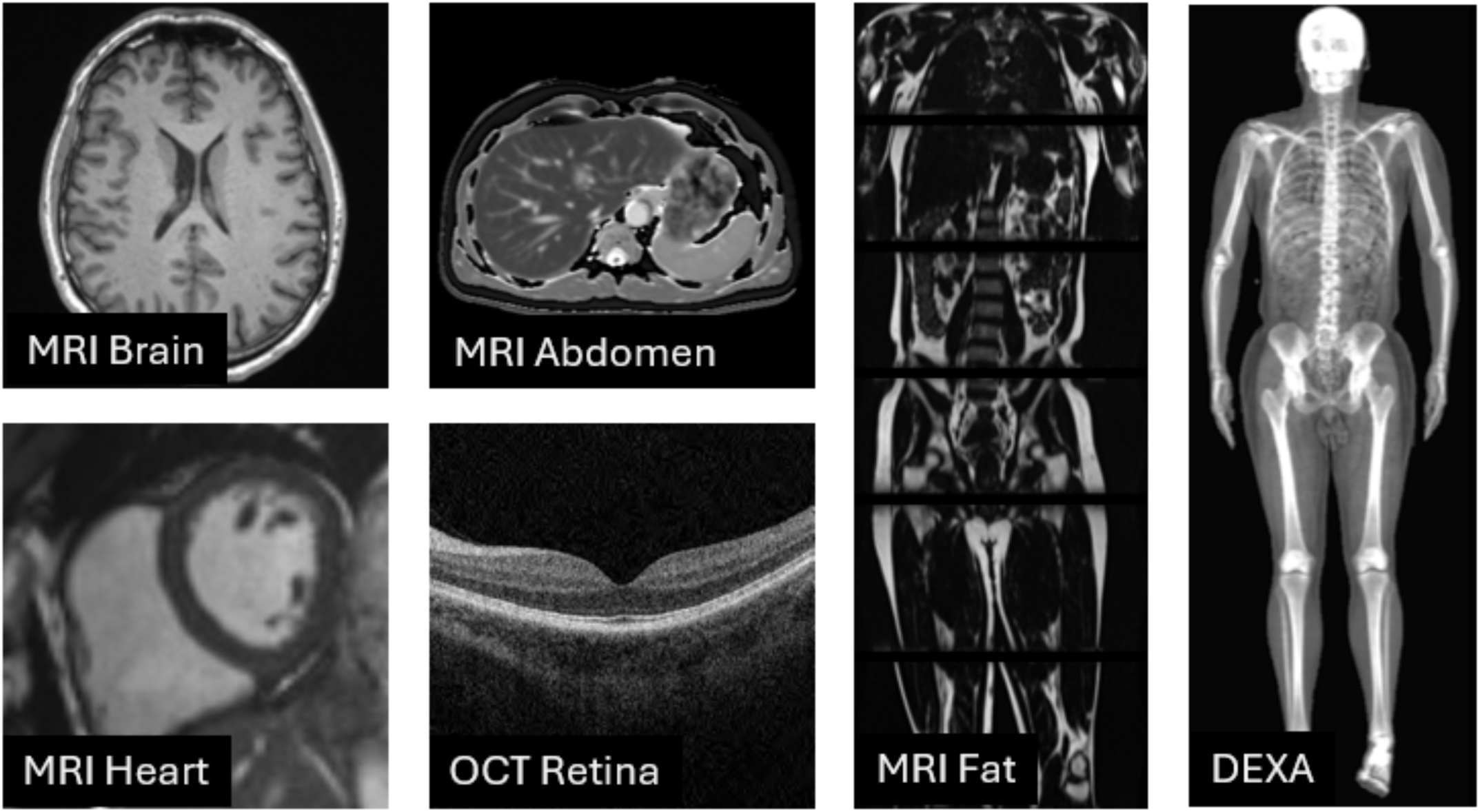
Example images from the UK Biobank Multi-organ Multimodal Imaging Assessment footnote. DEXA: Dual-energy X-ray Absorptiometry; MRI: magnetic resonance imaging; OCT: optical computed tomography

The UK Biobank is being continuously enhanced, leveraging emerging technologies to optimise its potential for scientific discovery and to address priority health problems of the time. Recent enrichments include whole genome sequencing, metabolomics analysis, and plasma proteomic profiling. A dedicated Brain Health Study is underway^22^, aiming to improve understanding of neurodegenerative conditions, such as Alzheimer’s dementia. The team are also piloting linkage to digital cancer histopathology images.

The UK Biobank provides non-preferential data access to researchers worldwide for approved health-related research^23^. This commitment to open science has been critical in allowing many researchers to work with the data in parallel and has resulted in the unique high impact of UK Biobank – with over 21,000 researchers across more than 60 countries working with the resource and more than 16,000 peer-reviewed scientific papers published as a result^24^.

### CMR imaging in UK Biobank

CMR examinations are performed on 1.5 Tesla scanners (MAGNETOM Aera, Syngo Platform VD13A, Siemens Healthcare, Erlangen, Germany) using a dedicated 20-minute acquisition protocol ^25,26^. Cine images are acquired using balanced steady state free precession sequences and include three long axis slices (two, three, and four-chamber), a short axis stack covering both ventricles from base to apex, a transverse view of the thoracic aorta, and sagittal and coronal views of the left ventricular outflow tract. Phase-contrast aortic flow sequences are acquired with velocity-encoding parameters manually adjusted to eliminate aliasing (starting at 2m/s).

The imaging protocol has been modified to reflect important developments in CMR post-processing technology. The UK Biobank initially included myocardial tagging sequences at three short axis levels (basal, mid-ventricular, apical). However, feature tracking strain, which can be derived from standard long and short axis cine images without any dedicated acquisitions has since become the established method for myocardial strain measurement. Myocardial tagging sequences were therefore dropped in July 2022. The scan time saved was used to extend native T1 mapping (Shortened Modified Look-Locker Inversion recovery – ShMOLLI, WIP780B) sequence) from one mid-ventricular slice to three short axis levels (basal, mid-ventricular, apical), allowing approximately 16-segment coverage of the left ventricle. Example images from the UK Biobank CMR protocol are provided in **Figure 2**.

**Figure 2.**
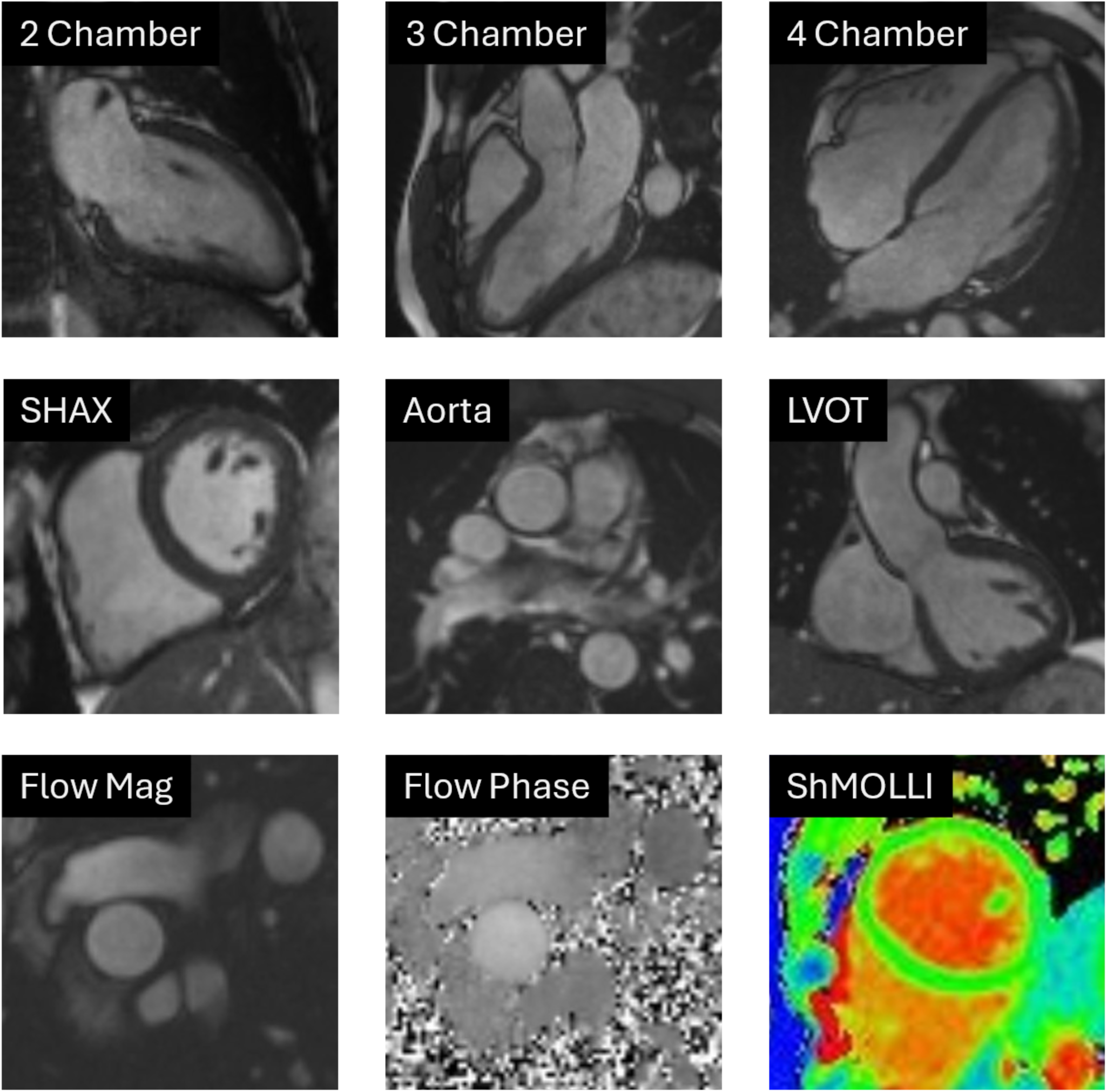
Example images from the UK Biobank Cardiovascular Magnetic Resonance Protocol footnote. Abbreviations: LVOT: left ventricular outflow tract; ShMOLLI: Modified Look-Locker Inversion recovery; SHAX: short axis.

Imaging is performed without administration of contrast or pharmacological stress agents, given the small (but present) risk of adverse reactions in the context of research scans performed for a very large sample in non-clinical settings^26^.

### Literature search

While our group has been responsible for setting up and guiding the UK Biobank CMR protocol during its lifetime so far, together with an international consensus group, a systemic literature review was also conducted to capture the different ways in which researchers have utilised UK Biobank CMR data. Google Scholar and Ovid Medline® (1946-January 2025) electronic databases were searched using pre-defined subject headings and key words and combined using Boolean operators. The “explode” function was applied to include the selected subject heading and all the more specific terms that are listed below it in the’tree’. An example of the search strategy is provided in **Table S1**. We included full length peer-reviewed original research publications from any discipline, written in the English language, that had used CMR data (whole images or derived phenotypes) from the UK Biobank. Works that did not directly use UK Biobank CMR data were not included, e.g. studies applying deep learning algorithms previously developed in UK Biobank on other datasets or those using only summary statistics from cardiovascular imaging genetic studies. Manuscripts in pre-print platforms, works only available as conference abstracts, brief reports, editorials, review papers, or research letters were not included. Full length original manuscripts published in proceedings of technical meetings – e.g., Institute of Electrical and Electronics Engineers (IEEE) were included.

After title and abstract screening, 463 papers were selected for full text review, and of these 282 met our inclusion criteria (**Table S2**). Articles were published between 2016 to 2025, with increasing number of publications year on year (**Figure 3)**. Publications originated across 19 countries, based on the primary institution of the senior author, with the largest number of papers from the United Kingdom (n=189), Unites States (n=37), and China (n=13) (**Figure 4**). The next sections discuss the most pertinent studies, nested in the overall context of how the UK Biobank has shaped the landscape of population science.

**Figure 3.**
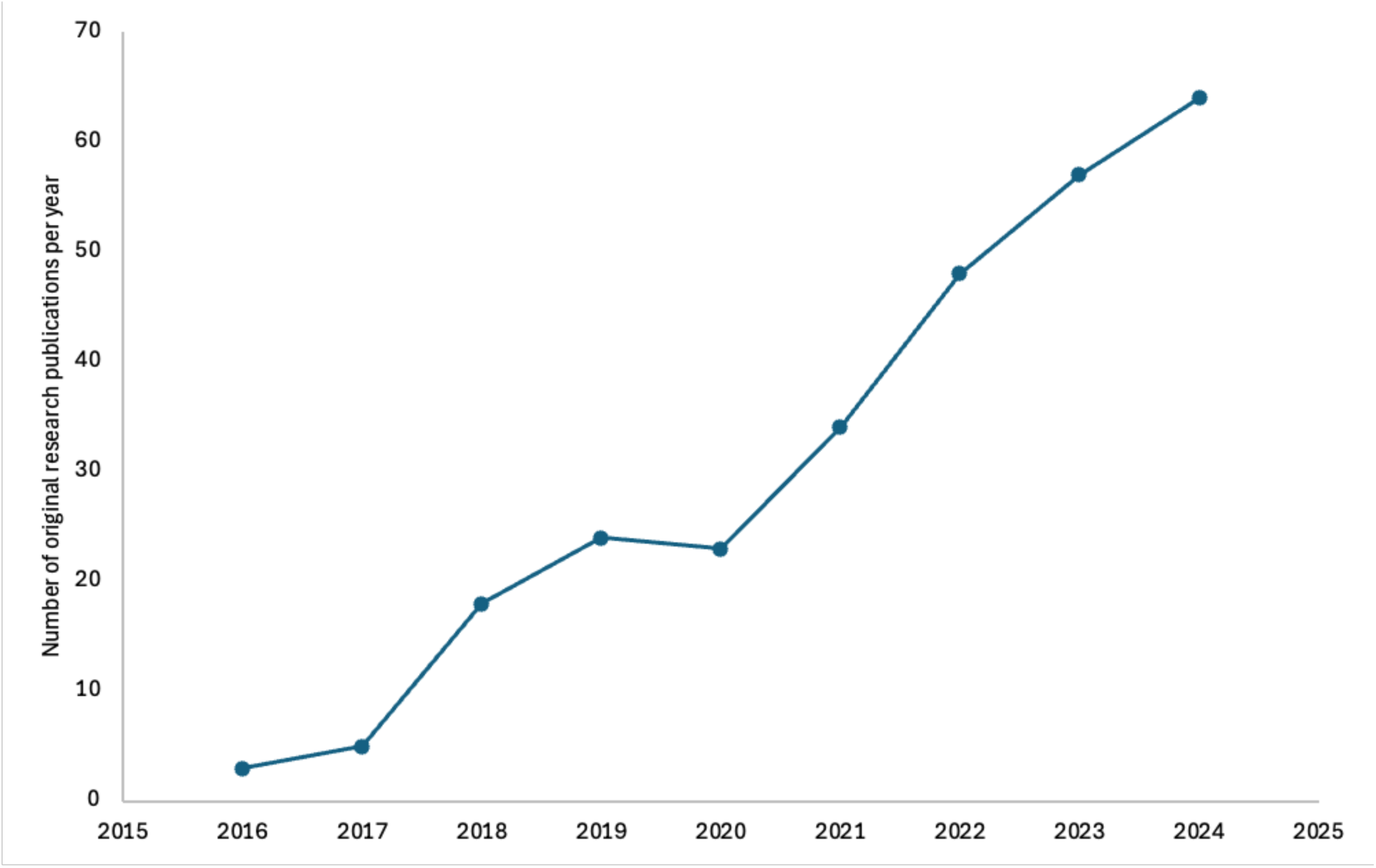
The number of original research publications using UK Biobank cardiovascular magnetic resonance data per year (showing data from 2016-2024)

### Large-scale image-based cardiovascular phenotyping

The UK Biobank CMR protocol allows capture of structure and function for all four cardiac chambers, aortic compliance, and myocardial tissue characterisation. The extraction of this information in the form of quantitative phenotypes requires quality control of input images, anatomic segmentation of cardiac structures, evaluation of segmentation quality, and parsing of the calculated metrics.

The acquisition of thousands of CMR scans in the UK Biobank necessitated development of automated and scalable pipelines for image analysis– a major challenge at the time. Towards this goal, the first 5,065 CMR scans were manually analysed in accordance with a pre-defined standard operating procedure across two core labs in Oxford and London (United Kingdom)^27^. This work generated a ground truth manual analysis dataset, which was used to develop and validate a fully automated quality-controlled convolutional neural network for extraction of volumetric CMR measures of cardiac structure and function^28^. In subsequent years, similar algorithms were developed for extraction of aortic strain and distensibility metrics^29^, global myocardial native T1 values^30,31^, and other less conventional phenotypes (e.g., mitral and tricuspid annular dimensions, mitral regurgitation detection)^32,33^. Artificial intelligence has since revolutionised the field of medical imaging, with fully/semi-automated cardiovascular image analysis tools commonly integrated (to varying degrees) in clinical and research settings – benefiting countless patients worldwide^34,35^.

The UK Biobank CMR dataset is now established as a key resource for driving technical innovations in cardiovascular imaging. Many researchers have used the UK Biobank to propose solutions for enhanced quality control of input images that may be automated and applied at scale. For example, detection of inadequate ventricular coverage^36–38^, suboptimal slice positioning or misalignment^39,40^, and detection (and sometimes correction) of incorrect gating^39^ or motion artefacts^41–45^. The resource has been used to develop methods that reduce the time and resources required to train robust segmentation models, through development of tools for automated image classification^46,47^, and demonstration of the role of synthetic data, weakly supervised deep learning, and model pre-training in reducing the size of manually labelled training samples required to achieve high model performance^48–50^. There has been substantial work on integrated automated approaches for quality control of generated segmentations^51–53^. End-to-end pipelines have been developed encompassing all these steps^54,55^.

The UK Biobank has also been used to investigate potential biases in automated image segmentation algorithms, with several researchers demonstrating no evidence of sex bias, but possible racial bias in performance of models trained on the resource, highlighting the importance of training samples with adequate representation of target populations^56–58^.

As well as extracting conventional CMR phenotypes, the UK Biobank has enabled large-scale investigation of novel imaging biomarkers such as radiomics features^59^, left ventricular trabeculations^60^, fractal dimensions^61^, and pericardial adiposity^62^. There have also been alternative approaches to quantifying the cardiovascular imaging phenotype using statistical cardiac atlases^63–65^ and cardiac shape models^66–68^. Some researchers have elected to work with whole image files (vs. derived metrics) and others have proposed to use raw data from K-space to extract more information than is perceptible from the derived whole images^69^.

The UK Biobank has also been instrumental in efforts to create high-quality virtual populations of human hearts, which is of major importance in a variety of applications, such as in silico simulations of cardiac physiology, data augmentation, and medical device development^70,71^.

### Cardiovascular Imaging Genomics

The UK Biobank CMR study provided high-resolution image-derived cardiovascular phenotypic data in large enough samples to allow reliable investigation of their genetic architecture – launching the field of cardiovascular imaging genomics, which aims to establish a) *heritability* of imaging phenotypes, b) *genetic variation* with imaging phenotypes to gain biological insights, and c) *personalised risk* prediction tools for clinical application.

Investigations may focus on candidate genes, selected a priori based on biologic understanding, which are usually rare variants with known causal links to rare conditions (e.g., HCM). The UK Biobank has enabled definition of the frequency of such rare variants in large populations and their associations with disease outcomes and imaging alterations^72–74^. Alternatively, genome wide association studies (GWASs) take the approach of “genetic discovery”, aiming to screen the whole genome to identify common genetic variants for common complex diseases. Functional mapping of identified variants can provide new insights into causal mechanisms, although linkage disequilibrium and inter-gene mapping present challenges to interpretation^75^. The genetic information from GWASs may be used to calculate polygenic risk scores estimating an individual’s genetic predisposition to a trait or disease.

Identified genetic variants may also be used as instrumental variables in Mendelian Randomisation analyses supporting causal inference^76^.

UK Biobank has enabled uncovering of the genetic architecture of cardiovascular imaging traits^77–79^, providing insight into the frequency and relevance of rare pathologic cardiomyopathy variants^73^, and evaluating the potential polygenicity of cardiomyopathies with previously understood monogenic inheritance^80^.

### Integration of imaging data into population science

There are many ways that researchers have integrated the UK Biobank CMR data into population science research – however, we can broadly think of these studies as having three structures with the imaging phenotypes considered as: a) *outcomes* used to understand the relationship of a genetic or environmental exposure with cardiovascular structure and function, b) *biomarkers* associated with future disease risk, and c) *mediators* used to delineate the potential mechanistic pathway through which a particular exposure leads to subsequent clinical disease.

Phenome-wide association studies have illustrated the value of population imaging in understanding the cardiovascular consequences of a wide range of environmental exposures^81^. Others have taken a more focused approach to evaluating specific exposures, providing new insights into the cardiovascular consequences of established risk factors and highlighting novel disease determinants. For instance, a detailed characterisation of the hypertensive cardiovascular phenotype in UK Biobank demonstrated associations with larger poorer functioning left atria, concentric left ventricular hypertrophy with reduced function, and reduced aortic compliance – and variations of these remodelling patterns by duration of hypertension exposure, blood pressure control, and across sex and ethnic groups^82^. Another study, reported the adverse impact of noise pollution, demonstrating association of higher nighttime aircraft noise levels with greater left ventricular mass and wall thickness, highlighting important considerations for urban planning and proximity of residential housing to airports^83^.

### Interpreting health and disease trends of CMR phenotypes in UK Biobank

Understanding trends of cardiovascular phenotypes in relation to health and disease is essential for integration of cardiovascular imaging in population research. This is not always straightforward.

Cardiovascular physiology and its compensatory adaptations to adverse exposures are complex and dynamic. The trajectory and direction of cardiovascular morpho-functional trends may be altered with exposure to stressors or onset of disease.

Previous CMR research cohorts had typically focused on smaller samples with high cardiometabolic disease burden. The UK Biobank provided the opportunity to define trends of cardiovascular phenotypes in a very large mostly healthy population, and to illustrate their modification with accumulation of risk factors and disease onset. The interpretation of CMR associations in this context requires special considerations and may present some pitfalls.

Cardiovascular phenotypes alter with healthy aging – comprising smaller cardiac chambers, lower left ventricular mass, more concentric left ventricular remodelling, higher ejection fraction, and lower global longitudinal strain^84^; an illustrative analysis of these age trends in the UK Biobank presented in **Figure 5**. In general, in the UK Biobank, the direction of association with unhealthy exposures follows that of greater cardiac aging (and healthy exposures show the reverse). This means that associations with larger cardiac chambers and lower ejection fraction (though broadly within normal ranges) may indicate healthy trends – the reverse of what we might expect from previous work in cohorts with greater disease burden.

**Figure 4.**
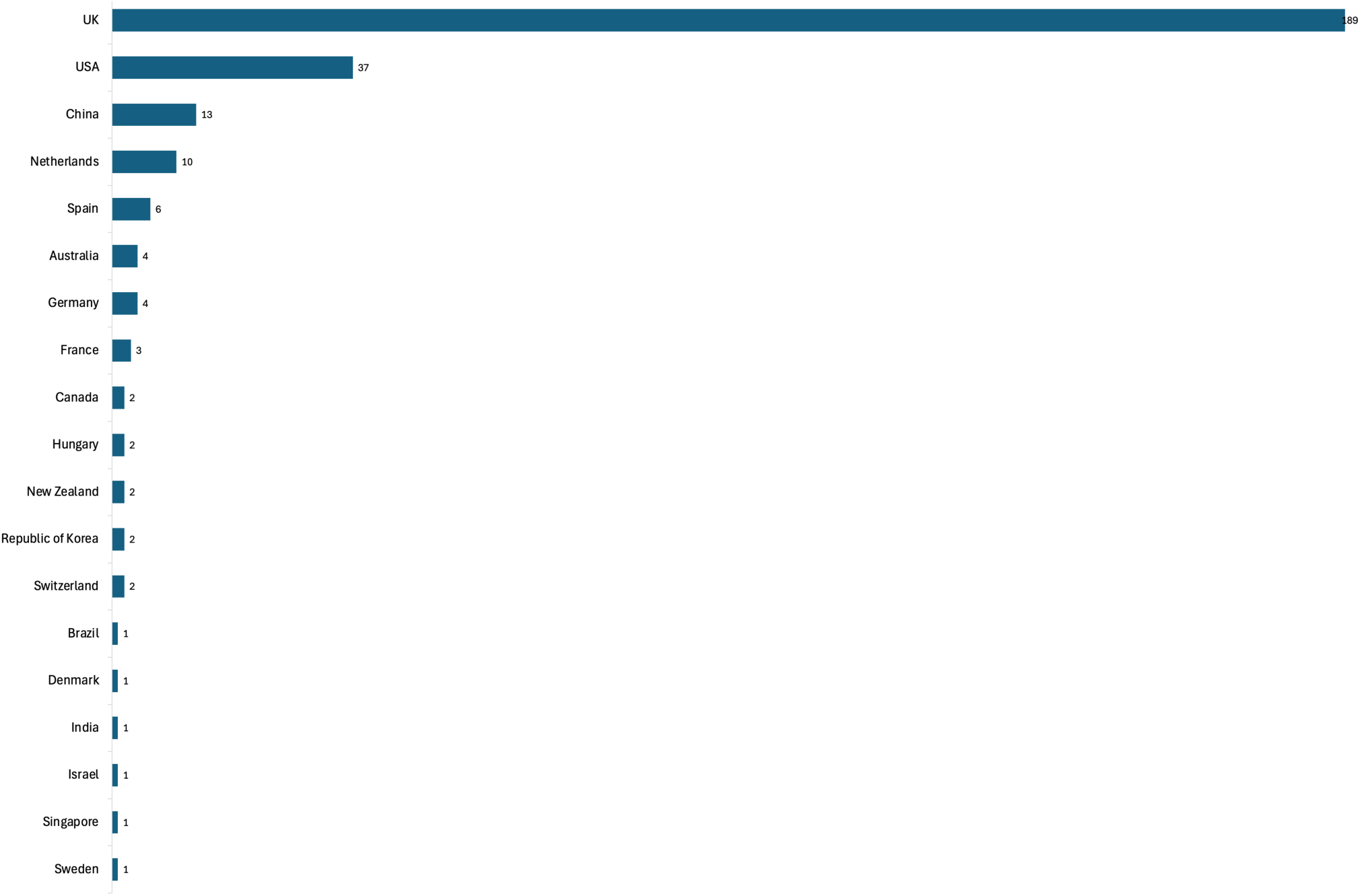
**The country of origin for original research publications using UK Biobank cardiovascular magnetic resonance data (based on the senior author’s main institution)**

**Figure 5.**
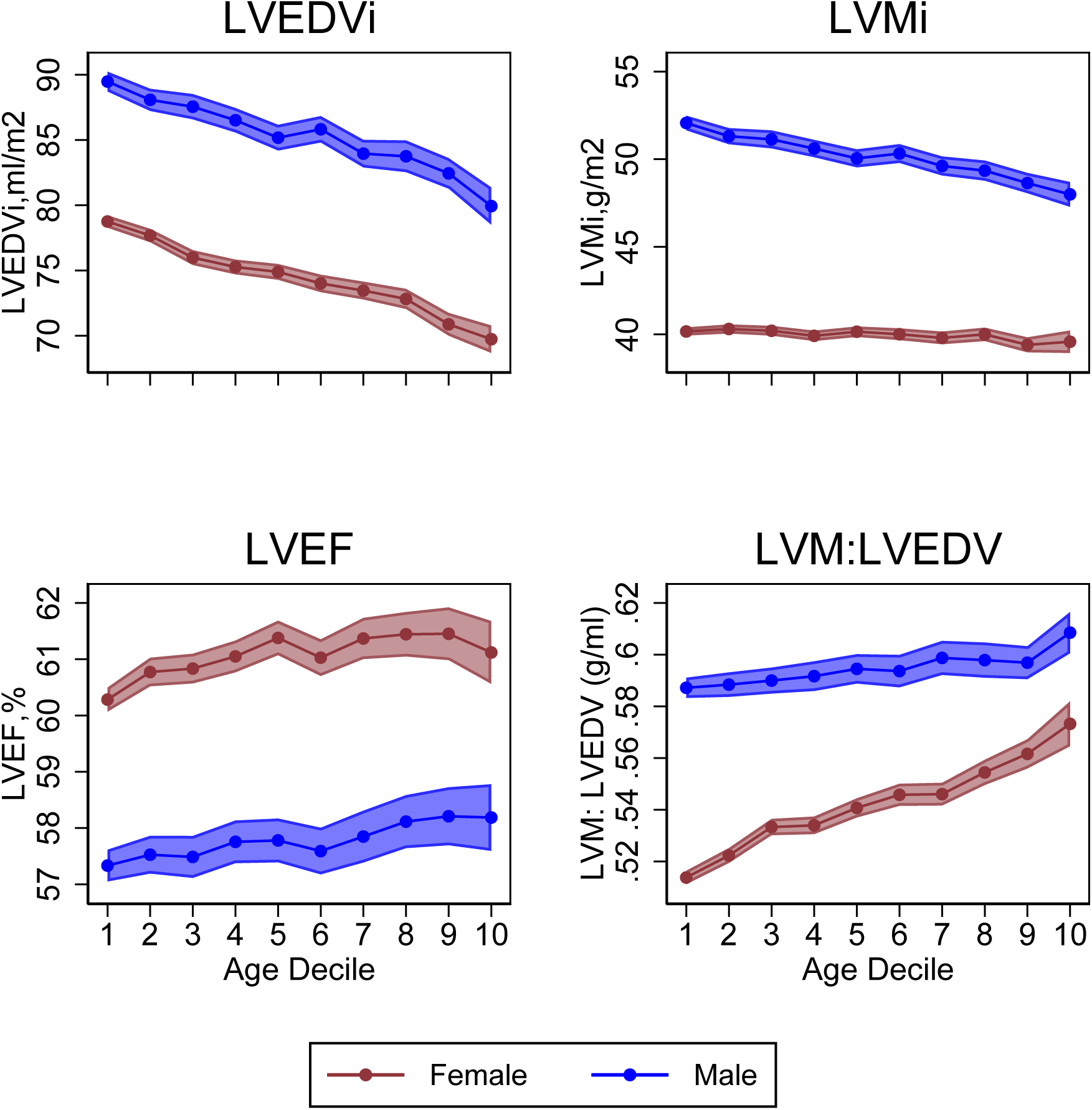
Cardiovascular magnetic resonance metrics in health stratified by age and sex footnote. The points represent mean cardiovascular magnetic resonance metrics and the shaded regions are 95% confidence intervals around the mean. Results are shown separately for women (red) and men (blue). The analysis included 18,744 UK Biobank participants with imaging data available and without history of cardiovascular disease at imaging. The sample is divided into approximately ten equal groups. Participants over the age of 80 are excluded due to small sample size. Abbreviations: LVEDVi: left ventricular end-diastolic volume indexed to body surface area; LVMi: left ventricular mass indexed to body surface area; LVEF: left ventricular ejection fraction; LVM: LVED: left ventricular mass to volume ratio.

The precise pattern of phenotypic alterations varies depending on the context. For example, in conditions where hypertrophy is a major part of the phenotypic manifestation (e.g., hypertension) we would expect, in the early disease stages, association with greater left ventricular mass but higher ejection fraction due to compensatory increase in radial function and concentric hypertrophy – in later stages, as compensatory adaptations begin to fail, the left ventricular cavity dilates (eccentric hypertrophy) and ejection fraction declines. The pattern of phenotypic associations with hypertension in a large cohort will depend on the average remodelling status within the sample, as well as the combined effect of all the other exposures that may be influencing cardiovascular adaptations.

Interpreting the trends of cardiovascular imaging phenotypes requires knowledge of cardiovascular physiology, consideration of the clinical context, and the overall pattern of phenotypic alterations (rather than relying on a single metric). There are some measures that have more consistent associations with health/disease and can be used as sense checks for interpretation of the overall picture – for instance, global longitudinal strain, aortic distensibility, and left ventricular global function index show more linear trends in response to adverse exposures and disease onset (**Figure 6**).

Understanding the disease distribution within the cohort and expected compensatory adaptations related to specific exposures are important for meaningful interpretation of trends of CMR metrics in large cohorts.

**Figure 6.**
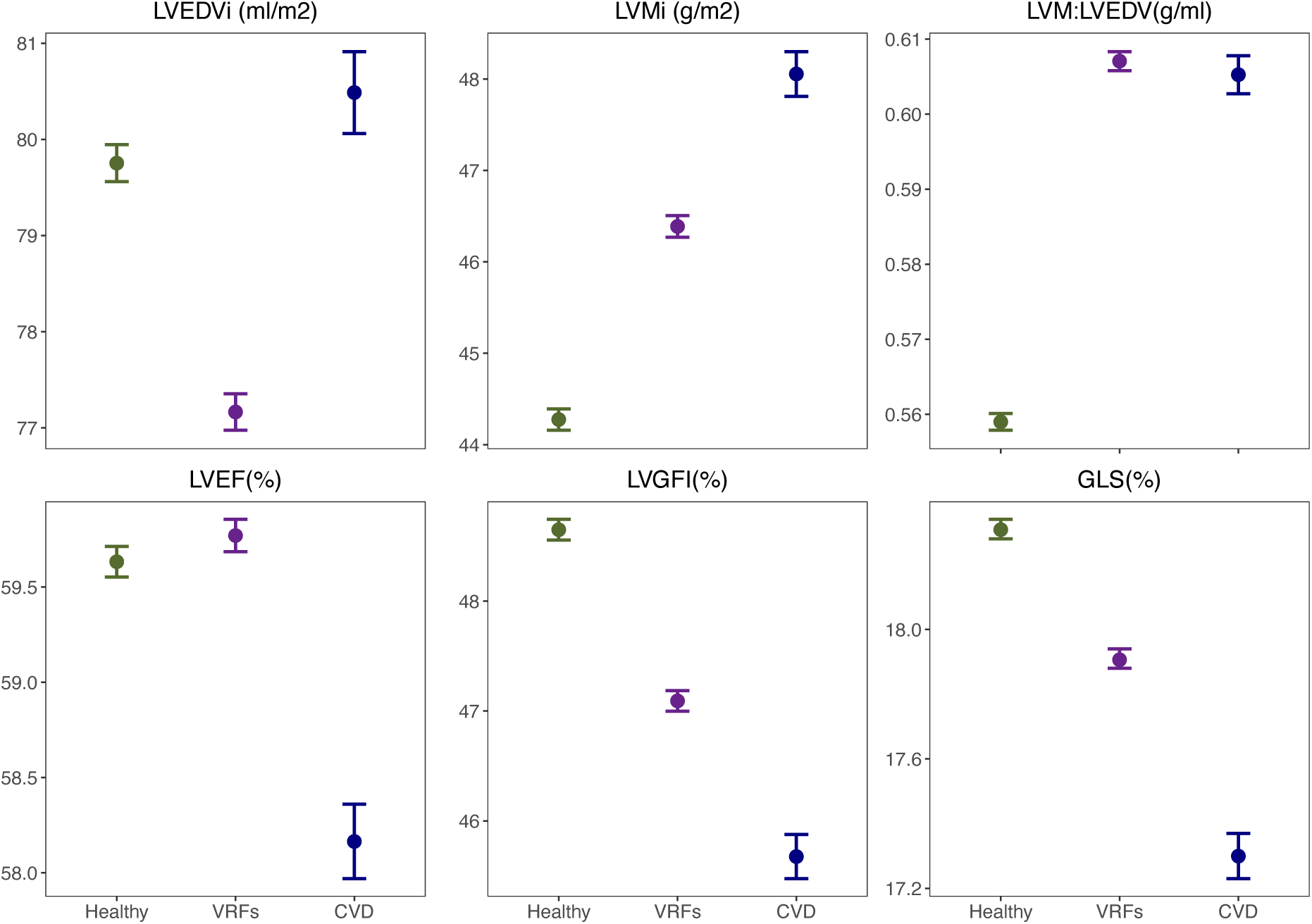
Cardiovascular metrics stratified by cardiovascular disease status footnote. The points indicate metric means, and the interval bars represent 95% confidence interval of the mean. The disease groups include healthy ( no VRFs, no CVDs), VRFs (prevalent VRFs, but no prevalent CVDs), CVDs (prevalent CVD with/without prevalent VRFs). The analysis sample included 45,349 UK Biobank participants with imaging data available. Abbreviations: CVD: cardiovascular disease; LVEDVi: left ventricular end-diastolic volume indexed to body surface area; LVEF: left ventricular ejection fraction; LVGFI: left ventricular global function index; GLS: global longitudinal strain; LVMi: left ventricular mass indexed to body surface area; LVM: LVED: left ventricular mass to volume ratio; VRF: vascular risk factor.

### Biological cardiovascular aging

The structural and functional alterations in cardiovascular aging are reliably captured on imaging and may be used to estimate a person’s “biological heart age”, which may differ from their chronological age^85^. The discrepancy between the biological and chronological age is termed the age “gap” or “delta” and may be considered an indicator of healthy (or unhealthy) cardiovascular aging. The UK Biobank has been used to model biological heart age using image-derived cardiovascular phenotypes, leveraged to map the relationship of environmental exposures with the heart age gap, provide new information about the genetic susceptibility to greater biological heart aging, and demonstrate associations of larger heart age gaps with greater risk of incident cardiovascular events^86–90^. This body of work has provided a new approach to phenotyping cardiovascular risk and has generated a comprehensive assessment of environmental and genetic determinants of biological aging of the human heart.

Following a similar concept, researchers built a model that learns not only what a human heart should look like but also how it should move during a heartbeat (based on thousands of CMRs), they then compared each individuals’ actual heart to that “normal” benchmark^91^. The degree of deviation was found to significantly associate with cardiovascular risk factors and disease, highlighting its potential utility for phenotyping subclinical dysfunction and enhancing predictive modelling.

### Distinction of health from disease

In clinical practice, deviation of CMR metrics from defined “healthy ranges” is used to guide diagnostic and treatment decisions. However, the healthy cardiovascular phenotype varies across different ages, sexes, ethnic groups, and among people of different body size^92^. Inadequate capture of these variations in clinical reference ranges may lead to misclassification of disease status.

The UK Biobank has been used to illustrate sex and ethnicity differences in cardiovascular structure and function metrics^27,93,94^. More recently, healthy reference ranges stratified by age, sex, ethnicity, and body size have been developed from the Healthy Hearts Consortium^95,96^, an international collaborative comprising the world’s largest CMR databank of verified healthy adults including contributions from the UK Biobank and five other research cohorts, which collectively capture the full adult age spectrum, adequate sex representation, and greater ethnic diversity than any previous resource – presenting a new international standard for defining the healthy adult heart with direct relevance to patient care.

Other researchers have focused on defining normal thresholds for CMR metrics of diagnostic significance within specific clinical contexts. For example, highlighting population variations of maximum wall thickness and the relevance of this to misclassification of HCM (where a fixed wall thickness threshold of ≥15mm is currently a diagnostic criteria) and proposing demographic-specific thresholds for more accurate disease discrimination^97,98^.

The UK Biobank has also permitted characterisation of population variations in select cardiovascular phenotypes of uncertain significance. For example, mitral annular disjunction, defined as a separation between the left atrial wall at the point of mitral valve insertion and the left ventricular free wall, has been linked to mitral valve disease and life-threatening arrhythmias^99^. However, understanding the distinct prognostic relevance of this anatomic variation is challenging, as it is almost always observed in clinical cohorts who have already experienced a serious adverse outcome or cardiac disease. A systematic evaluation in the UK Biobank found that this anatomic variation was overall very common and that only a less common subcategory (inferolateral disjunction) associated with mitral valve abnormalities – however, there were too few malignant arrhythmia events to exclude this association^100^; it may be possible to address this question in future years as more outcomes accrue.

Similarly, excess left ventricular trabeculation is a CMR phenotype previously linked to adverse outcomes (e.g., sudden death) in clinical cohorts with cardiomyopathy, with some proposing this phenotype as a distinct disease entity named “non-compaction cardiomyopathy”. Researchers have used the UK Biobank to characterise the variations of left ventricular trabeculation in health^60^, and its clinical and genetic associations^101^. The current expert consensus is that excess left ventricular trabeculation (in isolation) does not represent a disease or high-risk state, and that past reports of adverse associations in clinical cohorts reflect other co-existing cardiac abnormalities^102^. The investigations in UK Biobank contributed importantly to reaching this conclusion, which has benefitted many patients who were previously mislabelled with a serious cardiac condition.

### Imaging biomarkers for disease prediction

The availability of CMR in a large mostly healthy sample with longitudinal outcome tracking has provided a unique platform for evaluating the potential of imaging biomarkers for disease prediction.

Myocardial native T1 provides a non-invasive measure of tissue fibrosis with recognised prognostic value in clinical cohorts (e.g., in cardiomyopathies)^103^. Automated extraction of global myocardial T1 from the UK Biobank^30,31^ enabled illustration of its population variations and the first description of its value for predicting future cardiovascular events in people who did not have CVD at time of imaging^104^. Further work used myocardial T1 to uncover novel genetic loci related to greater myocardial interstitial fibrosis and provided new insights into potential biological pathways linking greater T1 values with CVD^105^.

Impaired myocardial relaxation or diastolic dysfunction is a sensitive indicator of CVD. Machine-learning cardiac motion analysis has been used to extract diastolic function metrics from UK Biobank cine CMR images, and to identify associations with key environmental determinants (e.g., diabetes), and elucidate novel associations with genetic traits related to maintaining sarcomeric function and genes implicated in the development of cardiomyopathy, and to demonstrate causally relevant relationships with heart failure^106^. Others have demonstrated the value of left atrial structure and function metrics (sensitive surrogate indicators of raised left ventricular filling pressure and diastolic dysfunction) in association with incident CVD events, beyond standard left ventricular metrics^107^.

The UK Biobank has also enabled the first large-scale analysis of the prognostic relevance of feature tracking strain, suggesting potential incremental clinical utility of these metrics for prediction of heart failure, myocardial infarction, and stroke^108^. This measurement can be extracted from standard-of-care images without the need for any dedicated acquisitions and using semi-automated clinical post-processing tools. The work in UK Biobank has contributed to a move towards adoption of these measurements in routine clinical practice.

Other novel imaging biomarkers with potential incremental predictive value over conventional CMR metrics (e.g., ventricular volume asymmetry and global longitudinal active strain energy density) have also been investigated with promising initial results^109,110^.

### Genetic determinants of cardiovascular imaging phenotypes

The UK Biobank has enabled characterisation of the genetic architecture of image-derived left and right ventricular structure and function, with functional mapping of identified variants providing new insights into susceptibility to heart failure and cardiomyopathies^77,78,111,112^.

The identification of novel loci associated with size and distensibility of the ascending aorta has provided insights into mechanisms underlying aortic size and function and provide genetic evidence for their role in aortic aneurysm development – a polygenic risk score based on this information has been proposed as potentially useful for guiding important clinical decisions such as need for surgical intervention^79,113,114^. Models integrating polygenic risk information and clinical risk factors have been developed with potential utility for detection of ascending aortic dilatation and adverse thoracic aortic events^115^. Others further demonstrate the potential role of genetic determinants of aortic strain and distensibility in influencing risk of stroke and coronary artery disease^116^, and suggest causal relevance of the relationships between aortic distensibility and cerebral microvascular injury^117^.

Understanding of the genetic aetiology of aortic valve disease has been advanced through discovery of genetic loci linked to aortic valve area and cardiac flow phenotypes, and demonstration of contributions from connective tissue genes, blood pressure, and root size to aortic valve function^118,119^. Similar analyses have been conducted in relation to the genetic basis of mitral valve phenotypes^120^.

Alternative approaches to image-based phenotyping have been taken with the aim of extracting new genetic information about determinants of cardiovascular morphometrics. For instance, through unsupervised feature extraction using autoencoders that operate on image-derived 3D meshes^121^ or by construction of cardiac shape metrics^122^.

### Novel insights into genetic cardiomyopathies

The imaging, genetic, and health outcome data in UK Biobank have been used to advance our understanding of the genetic architecture and phenotypic expression of hereditary cardiomyopathies and related structural cardiovascular traits.

An analysis of the population prevalence of rare sarcomere-encoding genes associated with HCM identified these variants in 2.9% of the UK Biobank cohort and demonstrated their association with an asymmetric increase in left ventricular maximum wall thickness and an increased risk of adverse cardiovascular outcomes, but with low aggregate penetrance to overt HCM^73^. Broadening the view to common genetic variation, two GWAS studies—one using deep learning to extract regional left ventricular wall thickness^80^ and another focused on maximum wall thickness^123^ identified numerous risk loci (e.g., 72 loci for regional thickness and shared polygenic signals with HCM). These polygenic determinants not only overlap with monogenic HCM genes but also stratify disease risk: higher polygenic scores predicted greater wall thickness and correlated with elevated HCM incidence. Complementing these, novel loci influencing septal structure were found to associate with both HCM and septal defects, underscoring shared developmental pathways in structural heart disease^124^. Finally, a data-driven taxonomy integrating genetics and imaging revealed HCM subgroups along a continuum of wall shape, genetic burden, and clinical outcomes, highlighting the interplay of rare monogenic and polygenic influences in shaping disease heterogeneity^125–127^.

Further studies have similarly illustrated the continuum of genetic influence on cardiac structure and function in the context of dilated cardiomyopathy. One study found that 1 of 6 participants with rare pathogenic dilated cardiomyopathy genetic variants exhibited early disease features, most commonly manifesting with arrhythmias in the absence of substantial ventricular dilation or dysfunction^128^. In a study of participants with stable coronary artery disease, those with rare pathogenic dilated cardiomyopathy variants had a 57% greater risk of death or major adverse cardiac events^129^. These carriers also had significantly lower left ventricular ejection fractions compared to non-carriers. A GWAS study of CMR traits in over 36,000 UK Biobank participants (with replication in MESA) identified 45 novel loci linked to cardiac structure and function, many near Mendelian cardiomyopathy genes^130^. A polygenic score for left ventricular end systolic volume was associated with incident dilated cardiomyopathy and modified cardiac phenotype even among TTN (titin) truncating variant carriers, highlighting the role of common variants in dilated cardiomyopathy pathogenesis^130^.

Another study evaluated the prevalence of disease-causing transthyretin (TTR) variants and evaluate associated phenotypes and outcomes, demonstrating that 1 in 1,000 UK Biobank participants carry pathogenic or likely pathogenic TTR variants (notably Val142Ile), with a significantly higher prevalence among individuals of African ancestry (∼4.3%) compared to those of European ancestry (∼0.02)^131^. These variant carriers exhibit increased left ventricular mass, prolonged PR intervals, and face elevated risks of heart failure and conduction disease. Despite the low overall rate of diagnosed amyloidosis (2.8%), these findings suggest possible clinical relevance of TTR variant carriers for early intervention and monitoring.

### New insights into cardiovascular disease determinants

While hypertension is established as a major cardiovascular risk factor, many gaps remain in risk stratification, treatment optimisation, and mechanistic understanding. A machine learning model was developed using 500 clinical and multi-organ imaging variables in the UK Biobank to longitudinally model development of high blood pressure, presenting a tool that could be used to identify people at risk of hypertension progression^132^. Imaging indices of cardiovascular haemodynamics have been used to evaluate the role of peripheral and systemic cardiovascular alterations in driving higher blood pressure^133^. Cardiovascular alterations related to hypertension were characterised in the UK Biobank, defining variations across sexes and ethnic groups, and with demonstration of the value of good blood pressure control in mitigating adverse cardiovascular remodelling^82^. These findings were extended to demonstrate their likely causal importance using Mendelian Randomisation to evaluate the association of systolic blood pressure with left ventricular mass^134^. Hypertension has multi-system end-organ consequences, which significantly determine prognostic outlook. With this consideration, a model was developed using simple clinical measures to capture end-organ effects of hypertensive disease defined using multi-organ imaging, presenting a method to identify high-risk patients with high burden of end-organ complications^135^.

The UK Biobank has also permitted characterisation of cardiovascular remodelling changes related to diabetes^136^. Other researchers have provided additional layers of granularity, by evaluating the associations of diabetes with CMR radiomics phenotypes^59^, characterising electrophysiological as well as cardiovascular imaging alterations related to the condition^137^, and considering the inter-relationships with obesity in determining subsequent cardiovascular risk^138^. The proteomics profiling in UK Biobank has been used to generate novel insights into the molecular mechanisms driving heart failure in diabetes, indicating a role for lower Erbb3 and higher Hspa2 levels by demonstrating their association with impaired contractility^139^.

The association of high cholesterol and serum lipids with cardiovascular phenotypes have been similarly outlined, demonstrating likely causal relationships between genetically predicted serum low-density lipoprotein, total cholesterol, and triglycerides and higher left ventricular mass^140,141^, while others have evaluated association of PCSK9 loss of function variants with cardiovascular imaging phenotypes and heart failure, demonstrating no significant association with either^142^.

The value of image-based body composition and obesity phenotyping in providing more informative characterisation of associated disease risk is increasingly recognised. The availability of multimodal obesity phenotyping in very large samples makes the UK Biobank a powerful resource for obesity research. The resource has been used to demonstrate obesity-related cardiovascular remodelling, the role of these alterations in mediating major CVDs, and the value of integrating body mass index and waist-to-hip ratio to better understand obesity-related cardiovascular risk^143^. The image-based approach to quantifying obesity available in UK Biobank, has allowed illustration of the importance of visceral abdominal obesity and liver fat deposition in driving adverse left ventricular remodelling over general (subcutaneous) obesity^144^, with possible differences across sexes^145^. Others additionally demonstrate the independent association of image-defined liver adipose deposition with adverse cardiovascular remodelling and association of liver T1 with cardiovascular outcomes^146–148^.

Growing research indicates a mechanistic role for metabolically active epicardial and pericardial adipose tissue (EPAT) in driving poorer cardiovascular health. The UK Biobank has permitted first evaluations of these relationships in large human samples. EPAT has been extracted from UK Biobank scans using automated algorithms, and linked to adverse lipid profile, poorer glycaemic control, systemic inflammation, and prevalent and incident CVDs, as well as remodelling changes suggestive of a heart failure with preserved ejection fraction phenotype^62,149,150^. Others have investigated potential associations with coronary artery disease^151^. GWASs have identified distinct loci associated with greater proportional pericardial adipose deposition and genetic instrumental variables have been used to investigate potential causality of observed associations^150,152^. Extraction of radiomics phenotypes from pericardial adipose area has been used to evaluate the importance of EPAT tissue character (defined using pixel-level signal-intensity based features) in associations with heart failure outcomes^153^.

The relationships between higher ambient air pollution and noise exposure have been linked to adverse cardiovascular remodelling^83,154^ – informing public health and urban planning strategies.

Cardiovascular associations of nutritional factors, including caffeine, alcohol, and meat consumption have also been evaluated across multiple studies^155–158^. The importance of mental health factors in cardiovascular risk has also been highlighted by studies showing associations of polygenic risk for schizophrenia and adverse childhood experiences with unhealthy cardiovascular remodelling^159,160^. The relevance of sleep quality and patterns has also been demonstrated^161,162^.

### Multisystem health

The extensive phenotyping and multiorgan imaging in UK Biobank have enabled “whole system” approaches to evaluating health and disease, and illustration of the inter-connectedness of major organ systems.

The interactions between respiratory^163^, renal^164^, liver^165^, and musculoskeletal^166–168^ health have been evaluated across multiple studies integrating imaging, spirometry, genetic, and health outcome data.

Multi-omics approaches have also been used to examine the heart-brain axis – illustrating relationships between better cognition and healthier cardiovascular phenotypes, genetic associations of likely causal importance between greater aortic distension and greater white matter hyperintensities, the role of shared vascular risk factors in driving disease across both organ systems, and the links between greater cardiac and brain aging and disease^169–176^.

Shared determinants of disease across multiple organ systems have been explored, for example by demonstrating the association of greater alcohol consumption with adverse cardiovascular remodelling, greater liver fat deposition, and reduced brain volumes extracted from multi-organ magnetic resonance images^158^. Others have highlighted shared genetic aetiology of cardiac dysfunction and dementia^177^.

Multiorgan imaging has been used to demonstrate that the relationships across the heart-liver-brain organ systems – indicating that pre-clinical organ-level alterations in one system are linked to adverse changes across the multiorgan network, independent of shared environmental risk factors^178^. Another study used machine learning to quantify interstitial fibrosis—via T1 mapping—in the liver, pancreas, heart, and kidneys of nearly 44,000 UK Biobank participants, linking elevated T1 times to multiple prevalent diseases and identifying 66 organ-specific genetic loci. Notably, individuals with high fibrosis in three or more organs had a threefold increase in mortality risk, underscoring shared pathological pathways and prognostic importance^179^.

## Conclusions

The UK Biobank CMR Study has transformed the landscape of cardiovascular research, setting a new standard for large-scale imaging studies. Its integration of high-quality imaging with genomics, clinical records, and lifestyle data has catalysed discoveries fostering inter-disciplinarity across multiple domains—engineering, epidemiology, and genomics—in ways previously unattainable. The model of equitable data access has democratised discovery, accelerating global research at a scale and pace unmatched by any other biomedical resource. The UK Biobank project to repeat imaging assessments in large subset of participants is currently underway and will enable longitudinal evaluation of interval change in organ-level health providing a unique window into population health, aging, and disease – which will mature into an invaluable resource in coming years.

### Future Perspectives

Looking ahead, the emphasis should shift toward harmonisation across multicohort studies, supporting broader generalisability and advancing clinical translation of findings, whether into patient care pathways or through biological validation. Comparisons across cohorts with different underlying populations, disease burden, and exposure profiles will strengthen reliability of observations and expand the range of research questions that may be addressed. Transparent, secure, and equitable data access procedures are essential to advance global scientific progress. Efforts to harmonise standards across research databases should be prioritised, aiming to facilitate responsible data sharing without compromising participant consent, privacy, or data security.

The sheer volume and accessibility of data also carry the risk of research waste, where uncoordinated “data mining” efforts result in publications that lack mechanistic insight or translational relevance.

Addressing this challenge will require a commitment to methodological rigour, awareness of selection, collider, and healthy participant biases, and a culture of multidisciplinary working with shared responsibility for generating meaningful, reproducible, and impactful science. The UK Biobank CMR experience offers a blueprint for future cohorts worldwide—not just in design and execution, but in fostering the skillsets and collaborative ethos needed to realise the full potential of population imaging science and improve human health.

## Data Availability

Data are available through the UK Biobank's data access procedures.

https://www.ukbiobank.ac.uk/use-our-data/apply-for-access/

---

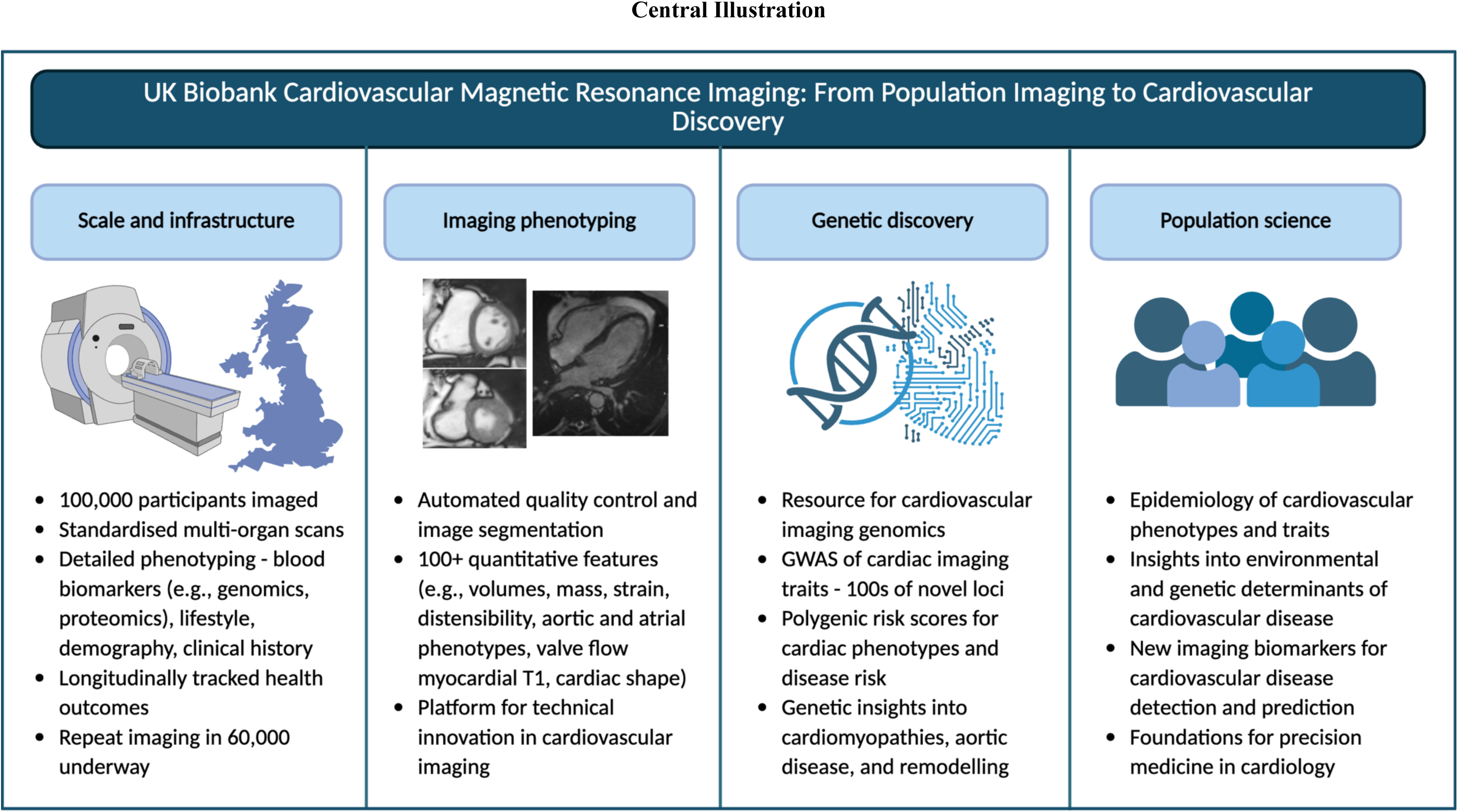

## SUPPLEMENTAL MATERIAL

**Table S1.** Example of search strategy performed on the Medline Ovid® electronic database.

| search | terms | results |
| --- | --- | --- |
| 1 | exp Magnetic Resonance Imaging/ or cardiovascular magnetic resonance.mp.<br>or exp Magnetic Resonance Imaging, Cine | 567472 |
| 2 | exp Cardiovascular System/ or exp Cardiovascular Abnormalities/ or<br>cardiovascular.mp. or exp Cardiovascular Diseases/ | 3777501 |
| 3 | heart.mp. or exp Heart/ | 1602414 |
| 4 | 2 or 3 | 4145385 |
| 5 | 1 and 4 | 125937 |
| 6 | CMR.mp. | 14639 |
| 7 | cardiovascular magnetic resonance.mp. | 7126 |
| 8 | 5 or 6 or 7 | 131783 |
| 9 | UK Biobank.mp. or exp UK Biobank/ | 10445 |
| 10 | 8 and 9 | 255 |

## Notes

### Author Declarations

Analysis of the UK Biobank was covered by the ethical approval for UK Biobank studies from the NHS National Research Ethics Service on 18th June 2021 (Ref 21/NW/0157) with written informed consent obtained from all participant. This work has been conducted using the UK Biobank application 2964.

## References

1. Ten Great Public Health Achievements - United States, 1900-1999. MMWR Morb Mortal Wkly Rep. 1999;48:241–243.

2. Achievements in Public Health, 1900-1999: Changes in the Public Health. J Am Med Assoc. 2000;283:735–738.

3. Mahmood SS, Levy D, Vasan RS, Wang TJ. The Framingham Heart Study and the epidemiology of cardiovascular disease: A historical perspective. Lancet 2014;383:999–1008.

4. Gaziano JM, Concato J, Brophy M, Fiore L, Pyarajan S, Breeling J, Whitbourne S, Deen J, Shannon C, Humphries D, Guarino P, Aslan M, Anderson D, Lafleur R, Hammond T, Schaa K, Moser J, Huang G, Muralidhar S, Przygodzki R, O’Leary TJ. Million Veteran Program: A mega-biobank to study genetic influences on health and disease. J Clin Epidemiol 2016;70:214–223.

5. Chen Z, Chen J, Collins R, Guo Y, Peto R, Wu F, Li L. China Kadoorie Biobank of 0.5 million people: survey methods, baseline characteristics and long-term follow-up. Int J Epidemiol 2011;40:1652–1666.

6. von Knobelsdorff-Brenkenhoff F, Schulz-Menger J. Cardiovascular magnetic resonance in the guidelines of the European Society of Cardiology: a comprehensive summary and update. J Cardiovasc Magn Reson 2023;25:42.

7. Messroghli DR, Moon JC, Ferreira VM, Grosse-Wortmann L, He T, Kellman P, Mascherbauer J, Nezafat R, Salerno M, Schelbert EB, Taylor AJ, Thompson R, Ugander M, Van Heeswijk RB, Friedrich MG. Clinical recommendations for cardiovascular magnetic resonance mapping of T1, T2, T2* and extracellular volume: A consensus statement by the Society for Cardiovascular Magnetic Resonance (SCMR) endorsed by the European Association for Cardiovascular Imaging (EACVI). J Cardiovasc Magn Reson 2017;19:1–24.

8. Lorenz CH, Walker ES, M VL, Kleit SS, Graham TP. Normal Human Right and Left Ventricular Mass, Systolic Function, and Gender Differences by Cine Magnetic Resonance Imaging. J Cardiovasc Magn Reson 1999;1:7–9.

9. Schroeder AP, Houlind K, Pedersen EM, Thuesen L, Nielsen TT, Egeblad H. Magnetic resonance imaging seems safe in patients with intracoronary stents. J Cardiovasc Magn Reson 2000;2:43–49.

10. Raman S V., Sparks EA, Baker PM, McCarthy B, Wooley CF. Mid-myocardial fibrosis by cardiac magnetic resonance in patients with lamin A/C cardiomyopathy: possible substrate for diastolic dysfunction. J Cardiovasc Magn Reson 2007;9:907–913.

11. Rutz AK, Juli CF, Ryf S, Widmer U, Kozerke S, Eckhardt BP, Boesiger P. Altered myocardial motion pattern in Fabry patients assessed with CMR-tagging. J Cardiovasc Magn Reson 2007;9:891–898.

12. Kramer CM, Appelbaum E, Desai MY, Desvigne-Nickens P, DiMarco JP, Friedrich MG, Geller N, Heckler S, Ho CY, Jerosch-Herold M, Ivey EA, Keleti J, Kim DY, Kolm P, Kwong RY, Maron MS, Schulz-Menger J, Piechnik S, Watkins H, Weintraub WS, Wu P, Neubauer S. Hypertrophic Cardiomyopathy Registry: The rationale and design of an international, observational study of hypertrophic cardiomyopathy. Am Heart J 2015;170:223–230.

13. Xu L, Pagano J, Chow K, Oudit GY, Haykowsky MJ, Mikami Y, Howarth AG, White JA, Howlett JG, Dyck JRB, Anderson TJ, Ezekowitz JA, Thompson RB, Paterson DI. Cardiac remodelling predicts outcome in patients with chronic heart failure. ESC Heart Fail 2021;8:5352–5362.

14. Miller RJH, Mikami Y, Heydari B, Wilton SB, James MT, Howarth AG, White JA, Lydell CP. Sex-specific relationships between patterns of ventricular remodelling and clinical outcomes. Eur Heart J Cardiovasc Imaging 2020;21:983–990.

15. Bild DE, Bluemke DA, Burke GL, Detrano R, Diez Roux A V., Folsom AR, Greenland P, Jacobs DR, Kronmal R, Liu K, Nelson JC, O’Leary D, Saad MF, Shea S, Szklo M, Tracy RP. Multi-Ethnic Study of Atherosclerosis: Objectives and Design. Am J Epidemiol 2002;156:871–881.

16. Bamberg F, Kauczor HU, Weckbach S, Schlett CL, Forsting M, Ladd SC, Greiser KH, Weber MA, Schulz-Menger J, Niendorf T, Pischon T, Caspers S, Amunts K, Berger K, Bülow R, Hosten N, Hegenscheid K, Kröncke T, Linseisen J, Günther M, Hirsch JG, Köhn A, Hendel T, Wichmann HE, Schmidt B, Jöckel KH, Hoffmann W, Kaaks R, Reiser MF, Völzke H. Whole-body MR imaging in the German national cohort: Rationale, design, and technical background1. Radiology 2015;277:206–220.

17. Littlejohns TJ, Holliday J, Gibson LM, Garratt S, Oesingmann N, Alfaro-Almagro F, Bell JD, Boultwood C, Collins R, Conroy MC, Crabtree N, Doherty N, Frangi AF, Harvey NC, Leeson P, Miller KL, Neubauer S, Petersen SE, Sellors J, Sheard S, Smith SM, Sudlow CLM, Matthews PM, Allen NE. The UK Biobank imaging enhancement of 100,000 participants: rationale, data collection, management and future directions. Nat Commun 2020;11:1–12.

18. Sudlow C, Gallacher J, Allen N, Beral V, Burton P, Danesh J, Downey P, Elliott P, Green J, Landray M, Liu B, Matthews P, Ong G, Pell J, Silman A, Young A, Sprosen T, Peakman T, Collins R. UK biobank: an open access resource for identifying the causes of a wide range of complex diseases of middle and old age. PLoS Med 2015;12.

19. Fry A, Littlejohns TJ, Sudlow C, Doherty N, Adamska L, Sprosen T, Collins R, Allen NE. Comparison of Sociodemographic and Health-Related Characteristics of UK Biobank Participants with Those of the General Population. Am J Epidemiol. 2017;186:1026–1034.

20. Petersen SE, Matthews PM, Bamberg F, Bluemke DA, Francis JM, Friedrich MG, Leeson P, Nagel E, Plein S, Rademakers FE, Young AA, Garratt S, Peakman T, Sellors J, Collins R, Neubauer S. Imaging in population science: cardiovascular magnetic resonance in 100,000 participants of UK Biobank - rationale, challenges and approaches. J Cardiovasc Magn Reson 2013;15:46.

21. Gibson LM, Nolan J, Littlejohns TJ, Mathieu E, Garratt S, Doherty N, Petersen S, Harvey NCW, Sellors J, Allen NE, Wardlaw JM, Jackson CA, Sudlow CLM. Factors associated with potentially serious incidental findings and with serious final diagnoses on multi-modal imaging in the UK Biobank Imaging Study: A prospective cohort study. PLoS One 2019;14.

22. The UK Biobank Brain Health Study - UK Biobank. [cited 2025 Jul 24];Available from: https://www.ukbiobank.ac.uk/the-uk-biobank-brain-health-study/

23. Allen NE, Sudlow C, Peakman T, Collins R. UK Biobank Data: Come and Get It. Sci Transl Med 2014;6:224ed4.

24. Use our data - UK Biobank. [cited 2025 Jul 24];Available from: https://www.ukbiobank.ac.uk/use-our-data/

25. Petersen SE, Matthews PM, Francis JM, Robson MD, Zemrak F, Boubertakh R, Young AA, Hudson S, Weale P, Garratt S, Collins R, Piechnik S, Neubauer S. UK Biobank’s cardiovascular magnetic resonance protocol. J Cardiovasc Magn Reson 2015;18:8.

26. UK Biobank Imaging modality Cardiovascular Magnetic Resonance (CMR). [cited 2025 Jul 10];Available from: http://www.ukbiobank.ac.uk/

27. Petersen SE, Aung N, Sanghvi MM, Zemrak F, Fung K, Paiva JM, Francis JM, Khanji MY, Lukaschuk E, Lee AM, Carapella V, Kim YJ, Leeson P, Piechnik SK, Neubauer S. Reference ranges for cardiac structure and function using cardiovascular magnetic resonance (CMR) in Caucasians from the UK Biobank population cohort. J Cardiovasc Magn Reson 2017;19:18.

28. Bai W, Sinclair M, Tarroni G, Oktay O, Rajchl M, Vaillant G, Lee AM, Aung N, Lukaschuk E, Sanghvi MM, Zemrak F, Fung K, Paiva JM, Carapella V, Kim YJ, Suzuki H, Kainz B, Matthews PM, Petersen SE, Piechnik SK, Neubauer S, Glocker B, Rueckert D. Automated cardiovascular magnetic resonance image analysis with fully convolutional networks. J Cardiovasc Magn Reson 2018;20:1–12.

29. Biasiolli L, Hann E, Lukaschuk E, Carapella V, Paiva JM, Aung N, Rayner JJ, Werys K, Fung K, Puchta H, Sanghvi MM, Moon NO, Thomson RJ, Thomas KE, Robson MD, Grau V, Petersen SE, Neubauer S, Piechnik SK. Automated localization and quality control of the aorta in cine CMR can significantly accelerate processing of the UK Biobank population data. PLoS One. 2019;14:e0212272.

30. Hann E, Popescu IA, Zhang Q, Gonzales RA, Barutçu A, Neubauer S, Ferreira VM, Piechnik SK. Deep neural network ensemble for on-the-fly quality control-driven segmentation of cardiac MRI T1 mapping. Med Image Anal 2021;71:102029.

31. Puyol-Antón E, Ruijsink B, Baumgartner CF, Masci P-G, Sinclair M, Konukoglu E, Razavi R, King AP. Automated quantification of myocardial tissue characteristics from native T1 mapping using neural networks with uncertainty-based quality-control. J Cardiovasc Magn Reson. 2020;22:60.

32. Gonzales RA, Lamy J, Seemann F, Heiberg E, Peters DC. Automated Measurements of Mitral and Tricuspid Annular Dimensions in Cardiovascular Magnetic Resonance. Proceedings - International Symposium on Biomedical Imaging. 2022;2022-March.

33. Xiao K, Learned-Miller E, Kalogerakis E, Priest J, Fiterau M. Machine Learning for Automated Mitral Regurgitation Detection from Cardiac Imaging. In: Greenspan, H., et al. Medical Image Computing and Computer Assisted Intervention – MICCAI 2023. MICCAI 2023. Lecture Notes in Computer Science, vol 14226. Springer, Cham. 10.1007/978-3-031-43990-2_23

34. Zhang Q, Fotaki A, Ghadimi S, Wang Y, Doneva M, Wetzl J, Delfino JG, O’Regan DP, Prieto C, Epstein FH. Improving the efficiency and accuracy of cardiovascular magnetic resonance with artificial intelligence-review of evidence and proposition of a roadmap to clinical translation. J Cardiovasc Magn Reson 2024;26.

35. Howard JP, Zhang Q, Salih A, Petersen SE, Lekadir K, Raisi-Estabragh Z. Artificial intelligence in cardiovascular imaging: risks, mitigations and the path to safe implementation. Heart. 2025;0:1–7.

36. Tarroni G, Oktay O, Bai W, Schuh A, Suzuki H, Passerat-Palmbach J, Glocker B, de Marvao A, O’Regan D, Cook S, Rueckert D. Learning-based heart coverage estimation for short-axis cine cardiac MR images. In: Pop M, Wright G. (eds) Functional Imaging and Modelling of the Heart. FIMH 2017. Lecture Notes in Computer Science(), vol 10263. Springer, Cham. 10.1007/978-3-319-59448-4_8

37. Zhang L, Pereañez M, Piechnik SK, Neubauer S, Petersen SE, Frangi AF. Multi-input and dataset-invariant adversarial learning (MDAL) for left and right-ventricular coverage estimation in cardiac MRI. In: Frangi A, Schnabel J, Davatzikos C, Alberola-López C, Fichtinger, G. (eds) Medical Image Computing and Computer Assisted Intervention – MICCAI 2018. MICCAI 2018. Lecture Notes in Computer Science(), vol 11071. Springer, Cham. 10.1007/978-3-030-00934-2_54

38. Zhang L, Gooya A, Pereanez M, Dong B, Piechnik SK, Neubauer S, Petersen SE, Frangi AF. Automatic Assessment of Full Left Ventricular Coverage in Cardiac Cine Magnetic Resonance Imaging with Fisher-Discriminative 3-D CNN. IEEE Trans Biomed Eng. 2019;66:1975–1986.

39. Banerjee A, Zacur E, Choudhury RP, Grau V. Optimised Misalignment Correction from Cine MR Slices Using Statistical Shape Model. In: Papież BW, Yaqub M, Jiao J, Namburete AIL, Noble JA. (eds) Medical Image Understanding and Analysis. MIUA 2021. Lecture Notes in Computer Science(), vol 12722. Springer, Cham. 10.1007/978-3-030-80432-9_16

40. Oksuz I, Ruijsink B, Puyol-Anton E, Sinclair M, Rueckert D, Schnabel JA, King AP. Automatic left ventricular outflow tract classification for accurate cardiac MR planning. Proceedings - International Symposium on Biomedical Imaging. 2018;2018-April:462–465.

41. Gonzales RA, Zhang Q, Papież BW, Werys K, Lukaschuk E, Popescu IA, Burrage MK, Shanmuganathan M, Ferreira VM, Piechnik SK. MOCOnet: Robust Motion Correction of Cardiovascular Magnetic Resonance T1 Mapping Using Convolutional Neural Networks. Front Cardiovasc Med 2021;8.

42. Tarroni G, Oktay O, Sinclair M, Bai W, Schuh A, Suzuki H, de Marvao A, O’Regan D, Cook S, Rueckert D. A comprehensive approach for learning-based fully-automated inter-slice motion correction for short-axis cine cardiac MR image stacks. In: Frangi A, Schnabel J, Davatzikos C, Alberola-López C, Fichtinger G. (eds) Medical Image Computing and Computer Assisted Intervention – MICCAI 2018. MICCAI 2018. Lecture Notes in Computer Science(), vol 11070. Springer, Cham. 10.1007/978-3-030-00928-1_31

43. Oksuz I, Ruijsink B, Puyol-Antón E, Bustin A, Cruz G, Prieto C, Rueckert D, Schnabel JA, King AP. Deep learning using K-space based data augmentation for automated cardiac MR motion artefact detection. In: Frangi A, Schnabel J, Davatzikos C, Alberola-López C, Fichtinger G. (eds) Medical Image Computing and Computer Assisted Intervention – MICCAI 2018. MICCAI 2018. Lecture Notes in Computer Science(), vol 11070. Springer, Cham. 10.1007/978-3-030-00928-1_29

44. Oksuz I, Clough J, Bustin A, Cruz G, Prieto C, Botnar R, Rueckert D, Schnabel JA, King AP. Cardiac MR motion artefact correction from k-space using deep learning-based reconstruction. In: Knoll, F., Maier, A., Rueckert, D. (eds) Machine Learning for Medical Image Reconstruction. MLMIR 2018. Lecture Notes in Computer Science(), vol 11074. Springer, Cham. 10.1007/978-3-030-00129-2_3

45. Sinclair M, Bai W, Puyol-Antón E, Oktay O, Rueckert D, King AP. Fully automated segmentation-based respiratory motion correction of multiplanar cardiac magnetic resonance images for large-scale datasets. In: Descoteaux, M., Maier-Hein, L., Franz, A., Jannin, P., Collins, D., Duchesne, S. (eds) Medical Image Computing and Computer-Assisted Intervention − MICCAI 2017. MICCAI 2017. Lecture Notes in Computer Science(), vol 10434. Springer, Cham. 10.1007/978-3-319-66185-8_38

46. Vergani V, Razavi R, Puyol-Antón E, Ruijsink B. Deep Learning for Classification and Selection of Cine CMR Images to Achieve Fully Automated Quality-Controlled CMR Analysis From Scanner to Report. Front Cardiovasc Med 2021;8.

47. Vergani V, Razavi R, Puyol-Antón E, Ruijsink B. Deep Learning for Classification and Selection of Cine CMR Images to Achieve Fully Automated Quality-Controlled CMR Analysis From Scanner to Report. Front Cardiovasc Med 2021;8.

48. Daum D, Osuala R, Riess A, Kaissis G, Schnabel JA, Di Folco M. On Differentially Private 3D Medical Image Synthesis with Controllable Latent Diffusion Models. In: Mukhopadhyay, A., Oksuz, I., Engelhardt, S., Mehrof, D., Yuan, Y. (eds) Deep Generative Models. DGM4MICCAI 2024. Lecture Notes in Computer Science, vol 15224. Springer, Cham. 10.1007/978-3-031-72744-3_14

49. Fries JA, Varma P, Chen VS, Xiao K, Tejeda H, Saha P, Dunnmon J, Chubb H, Maskatia S, Fiterau M, Delp S, Ashley E, Ré C, Priest JR. Weakly supervised classification of aortic valve malformations using unlabeled cardiac MRI sequences. Nat Commun 2019;10.

50. Gheorghiță BA, Itu LM, Sharma P, Suciu C, Wetzl J, Geppert C, Ali MAA, Lee AM, Piechnik SK, Neubauer S, Petersen SE, Schulz-Menger J, Chițiboi T. Improving robustness of automatic cardiac function quantification from cine magnetic resonance imaging using synthetic image data. Sci Rep 2022;12.

51. Ng M, Guo F, Biswas L, Petersen SE, Piechnik SK, Neubauer S, Wright G. Estimating Uncertainty in Neural Networks for Cardiac MRI Segmentation: A Benchmark Study. IEEE Trans Biomed Eng 2023;70:1955–1966.

52. Robinson R, Valindria V V., Bai W, Oktay O, Kainz B, Suzuki H, Sanghvi MM, Aung N, Paiva JM, Zemrak F, Fung K, Lukaschuk E, Lee AM, Carapella V, Kim YJ, Piechnik SK, Neubauer S, Petersen SE, Page C, Matthews PM, Rueckert D, Glocker B. Automated quality control in image segmentation: Application to the UK Biobank cardiovascular magnetic resonance imaging study. J Cardiovasc Magn Reson 2019;21.

53. Hann E, Biasiolli L, Zhang Q, Popescu IA, Werys K, Lukaschuk E, Carapella V, Paiva JM, Aung N, Rayner JJ, Fung K, Puchta H, Sanghvi MM, Moon NO, Thomas KE, Ferreira VM, Petersen SE, Neubauer S, Piechnik SK. Quality Control-Driven Image Segmentation Towards Reliable Automatic Image Analysis in Large-Scale Cardiovascular Magnetic Resonance Aortic Cine Imaging. In: Shen, D., et al. Medical Image Computing and Computer Assisted Intervention – MICCAI 2019. MICCAI 2019. Lecture Notes in Computer Science(), vol 11765. Springer, Cham. 10.1007/978-3-030-32245-8_83

54. MacHado I, Puyol-Anton E, Hammernik K, Cruz G, Ugurlu D, Olakorede I, Oksuz I, Ruijsink B, Castelo-Branco M, Young A, Prieto C, Schnabel J, King A. A Deep Learning-Based Integrated Framework for Quality-Aware Undersampled Cine Cardiac MRI Reconstruction and Analysis. IEEE Trans Biomed Eng 2024;71:855–865.

55. Attar R, Pereañez M, Gooya A, Albà X, Zhang L, de Vila MH, Lee AM, Aung N, Lukaschuk E, Sanghvi MM, Fung K, Paiva JM, Piechnik SK, Neubauer S, Petersen SE, Frangi AF. Quantitative CMR population imaging on 20,000 subjects of the UK Biobank imaging study: LV/RV quantification pipeline and its evaluation. Med Image Anal 2019;56:26–42.

56. Puyol-Antón E, Ruijsink B, Piechnik SK, Neubauer S, Petersen SE, Razavi R, King AP. Fairness in Cardiac MR Image Analysis: An Investigation of Bias Due to Data Imbalance in Deep Learning Based Segmentation. In: de Bruijne, M., et al. Medical Image Computing and Computer Assisted Intervention – MICCAI 2021. MICCAI 2021. Lecture Notes in Computer Science(), vol 12903. Springer, Cham. 10.1007/978-3-030-87199-4_39

57. Lee T, Puyol-Antón E, Ruijsink B, Shi M, King AP. A Systematic Study of Race and Sex Bias in CNN-Based Cardiac MR Segmentation. In: Camara, O., et al. Statistical Atlases and Computational Models of the Heart. Regular and CMRxMotion Challenge Papers. STACOM 2022. Lecture Notes in Computer Science, vol 13593. Springer, Cham. 10.1007/978-3-031-23443-9_22

58. Lee T, Puyol-Antón E, Ruijsink B, Aitcheson K, Shi M, King AP. An Investigation into the Impact of Deep Learning Model Choice on Sex and Race Bias in Cardiac MR Segmentation. In: Wesarg, S., et al. Clinical Image-Based Procedures, Fairness of AI in Medical Imaging, and Ethical and Philosophical Issues in Medical Imaging. CLIP EPIMI FAIMI 2023 2023 2023. Lecture Notes in Computer Science, vol 14242. Springer, Cham. 10.1007/978-3-031-45249-9_21

59. Raisi-Estabragh Z, Jaggi A, Gkontra P, McCracken C, Aung N, Munroe PB, Neubauer S, Harvey NC, Lekadir K, Petersen SE. Cardiac Magnetic Resonance Radiomics Reveal Differential Impact of Sex, Age, and Vascular Risk Factors on Cardiac Structure and Myocardial Tissue. Front Cardiovasc Med. 2021;8:1972.

60. Aung N, Bartoli A, Rauseo E, Cortaredona S, Sanghvi MM, Fournel J, Ghattas B, Khanji MY, Petersen SE, Jacquier A. Left Ventricular Trabeculations at Cardiac MRI: Reference Ranges and Association with Cardiovascular Risk Factors in UK Biobank. Radiology 2024;311.

61. Meyer H V., Dawes TJW, Serrani M, Bai W, Tokarczuk P, Cai J, de Marvao A, Henry A, Lumbers RT, Gierten J, Thumberger T, Wittbrodt J, Ware JS, Rueckert D, Matthews PM, Prasad SK, Costantino ML, Cook SA, Birney E, O’Regan DP. Genetic and functional insights into the fractal structure of the heart. Nature 2020;584:589–594.

62. Bard A, Raisi-Estabragh Z, Ardissino M, Lee AM, Pugliese F, Dey D, Sarkar S, Munroe PB, Neubauer S, Harvey NC, Petersen SE. Automated Quality-Controlled Cardiovascular Magnetic Resonance Pericardial Fat Quantification Using a Convolutional Neural Network in the UK Biobank. Front Cardiovasc Med 2021;8:1–11.

63. Puyol-Antón E, Sinclair M, Gerber B, Amzulescu MS, Langet H, Craene M De, Aljabar P, Piro P, King AP. A multimodal spatiotemporal cardiac motion atlas from MR and ultrasound data. Med Image Anal 2017;40:96–110.

64. Mauger C, Gilbert K, Lee AM, Sanghvi MM, Aung N, Fung K, Carapella V, Piechnik SK, Neubauer S, Petersen SE, Suinesiaputra A, Young AA. Right ventricular shape and function: Cardiovascular magnetic resonance reference morphology and biventricular risk factor morphometrics in UK Biobank. J Cardiovasc Magn Reson 2019;21.

65. Gilbert K, Suinesiaputra A, Neubauer S, Piechnik S, Aung N, Petersen SE, Young A. End-Diastolic and End-Systolic LV Morphology in the Presence of Cardiovascular Risk Factors: A UK Biobank Study. In: Coudière, Y., Ozenne, V., Vigmond, E., Zemzemi, N. (eds) Functional Imaging and Modeling of the Heart. FIMH 2019. Lecture Notes in Computer Science(), vol 11504. Springer, Cham. 10.1007/978-3-030-21949-9_33

66. Bonazzola R, Ferrante E, Ravikumar N, Xia Y, Keavney B, Plein S, Syeda-Mahmood T, Frangi AF. Unsupervised ensemble-based phenotyping enhances discoverability of genes related to left-ventricular morphology. Nat Mach Intell 2024;6:291–306.

67. Beetz M, Yang Y, Banerjee A, Li L, Grau V. 3D Shape-Based Myocardial Infarction Prediction Using Point Cloud Classification Networks. *Proceedings of the Annual International Conference of the IEEE Engineering in Medicine and Biology Society*, EMBS. 2023;

68. Zakeri A, Hokmabadi A, Ravikumar N, Frangi AF, Gooya A. A probabilistic deep motion model for unsupervised cardiac shape anomaly assessment. Med Image Anal 2022;75.

69. Li R, Pan J, Zhu Y, Ni J, Rueckert D. Classification, Regression and Segmentation Directly from K-Space in Cardiac MRI. In: Xu, X., Cui, Z., Rekik, I., Ouyang, X., Sun, K. (eds) Machine Learning in Medical Imaging. MLMI 2024. Lecture Notes in Computer Science, vol 15241. Springer, Cham. 10.1007/978-3-031-73284-3_4

70. Peng J, Beetz M, Banerjee A, Chen M, Grau V. Generating Virtual Populations of 3D Cardiac Anatomies with Snowflake-Net. In: Camara, O., et al. Statistical Atlases and Computational Models of the Heart. Regular and CMRxRecon Challenge Papers. STACOM 2023. Lecture Notes in Computer Science, vol 14507. Springer, Cham. 10.1007/978-3-031-52448-6_16

71. Kalaie S, Bulpitt A, Frangi AF, Gooya A, Kalaie S, Bulpitt A, Frangi A, Gooya A. A Geometric Deep Learning Framework for Generation of Virtual Left Ventricles as Graphs Proc Mach Learn Res 2024;227:426–443. Available from: https://proceedings.mlr.press/v227/kalaie24a.html

72. Rämö JT, Jurgens SJ, Kany S, Choi SH, Wang X, Smirnov AN, Friedman SF, Maddah M, Khurshid S, Ellinor PT, Pirruccello JP. Rare Genetic Variants in LDLR, APOB, and PCSK9 Are Associated with Aortic Stenosis. Circulation 2024;150:1767–1780.

73. de Marvao A, McGurk KA, Zheng SL, Thanaj M, Bai W, Duan J, Biffi C, Mazzarotto F, Statton B, Dawes TJW, Savioli N, Halliday BP, Xu X, Buchan RJ, Baksi AJ, Quinlan M, Tokarczuk P, Tayal U, Francis C, Whiffin N, Theotokis PI, Zhang X, Jang M, Berry A, Pantazis A, Barton PJR, Rueckert D, Prasad SK, Walsh R, Ho CY, Cook SA, Ware JS, O’Regan DP. Phenotypic Expression and Outcomes in Individuals With Rare Genetic Variants of Hypertrophic Cardiomyopathy. J Am Coll Cardiol 2021;78:1097–1110.

74. Jones RE, Hammersley DJ, Zheng S, McGurk KA, de Marvao A, Theotokis PI, Owen R, Tayal U, Rea G, Hatipoglu S, Buchan RJ, Mach L, Curran L, Lota AS, Simard F, Reddy RK, Talukder S, Yoon WY, Vazir A, Pennell DJ, O’Regan DP, Baksi AJ, Halliday BP, Ware JS, Prasad SK. Assessing the association between genetic and phenotypic features of dilated cardiomyopathy and outcome in patients with coronary artery disease. Eur J Heart Fail 2024;26:46–55.

75. Nayor M, Shen L, Hunninghake GM, Kochunov P, Barr RG, Bluemke DA, Broeckel U, Caravan P, Cheng S, De Vries PS, Hoffmann U, Kolossváry M, Li H, Luo J, McNally EM, Thanassoulis G, Arnett DK, Vasan RS. Progress and Research Priorities in Imaging Genomics for Heart and Lung Disease: Summary of an NHLBI Workshop. Circ Cardiovasc Imaging 2021;14:E012943.

76. Sanderson E, Glymour MM, Holmes M V., Kang H, Morrison J, Munafò MR, Palmer T, Schooling CM, Wallace C, Zhao Q, Davey Smith G. Mendelian randomization. Nature Reviews Methods Primers 2022;2:1–21.

77. Pirruccello JP, Di Achille P, Nauffal V, Nekoui M, Friedman SF, Klarqvist MDR, Chaffin MD, Weng LC, Cunningham JW, Khurshid S, Roselli C, Lin H, Koyama S, Ito K, Kamatani Y, Komuro I, Matsuda K, Yamanashi Y, Furukawa Y, Morisaki T, Murakami Y, Kamatani Y, Mutu K, Nagai A, Obara W, Yamaji K, Takahashi K, Asai S, Takahashi Y, Suzuki T, Sinozaki N, Yamaguchi H, Minami S, Murayama S, Yoshimori K, Nagayama S, Obata D, Higashiyama M, Masumoto A, Koretsune Y, Jurgens SJ, Benjamin EJ, Batra P, Natarajan P, Ng K, Hoffmann U, Lubitz SA, Ho JE, Lindsay ME, Philippakis AA, Ellinor PT. Genetic analysis of right heart structure and function in 40,000 people. Nat Genet 2022;54:792–803.

78. Aung N, Vargas JD, Yang C, Cabrera CP, Warren HR, Fung K, Tzanis E, Barnes MR, Rotter JI, Taylor KD, Manichaikul AW, Lima JAC, Bluemke DA, Piechnik SK, Neubauer S, Munroe PB, Petersen SE. Genome-wide analysis of left ventricular image-derived phenotypes identifies fourteen loci associated with cardiac morphogenesis and heart failure development. Circulation 2019;140:1318–1330.

79. Tcheandjieu C, Xiao K, Tejeda H, Lynch JA, Ruotsalainen S, Bellomo T, Palnati M, Judy R, Klarin D, Kember RL, Verma S, Mitnaul LJ, Mighty J, Jones MB, Ziyatdinov A, Zhang B, Ye B, Watanabe K, Stahl E, Sidore C, Paulding C, Moscati A, Mbatchou J, Marcketta A, Marchini J, Locke A, Liu D, Lin N, Li A, Kosmicki J, Kessler M, Kang HM, Jorgenson E, Gurski L, Gillies C, Ghosh A, Ferreira MAR, Dobbyn L, Damask A, Backman J, Xin J, Sun K, Salerno W, Rasool A, Polanco T, Panea R, Orelus M, O’Keeffe S, Nafde M, Mitra G, Maxwell EK, Mansfield AJ, Lanche R, Krasheninina O, Khalid S, Hawes A, Habegger L, Eom G, Chen S, Boutkov B, Bao S, Balasubramanian S, Bai X, Staples JC, Sharma D, Malhotra S, Li D, Banerjee N, Averitt A, Ulloa RH, Wolf SE, Widom L, Schleicher TD, Manoochehri K, Pradhan M, Padilla MS, Lopez A, Lattari M, Gu Z, Fuller ED, Forsythe C, Beechert C, Shuldiner A, Siminovitch K, Reid JG, Overton JD, Lotta LA, Karalis K, Economides A, Deubler A, Coppola G, Cantor M, Baras A, Abecasis G, Palotie A, Daly M, Ritchie M, Rader DJ, et al. High heritability of ascending aortic diameter and trans-ancestry prediction of thoracic aortic disease. Nat Genet 2022;54:772–782.

80. Ning C, Fan L, Jin M, Wang W, Hu Z, Cai Y, Chen L, Lu Z, Zhang M, Chen C, Li Y, Zhang F, Wang W, Liu Y, Chen S, Jiang Y, He C, Wang Z, Chen X, Li H, Li G, Ma Q, Geng H, Tian W, Zhang H, Liu B, Xia Q, Yang X, Liu Z, Li B, Zhu Y, Li X, Zhang S, Tian J, Miao X. Genome-wide association analysis of left ventricular imaging-derived phenotypes identifies 72 risk loci and yields genetic insights into hypertrophic cardiomyopathy. Nat Commun 2023;14:1–15.

81. Bai W, Suzuki H, Huang J, Francis C, Wang S, Tarroni G, Guitton F, Aung N, Fung K, Petersen SE, Piechnik SK, Neubauer S, Evangelou E, Dehghan A, O’Regan DP, Wilkins MR, Guo Y, Matthews PM, Rueckert D. A population-based phenome-wide association study of cardiac and aortic structure and function. Nat Med 2020;26:1654–1662.

82. Elghazaly H, McCracken C, Szabo L, Malcolmson J, Manisty CH, Davies AH, Piechnik SK, Harvey NC, Neubauer S, Mohiddin SA, Petersen SE, Raisi-Estabragh Z. Characterizing the hypertensive cardiovascular phenotype in the UK Biobank. Eur Heart J Cardiovasc Imaging 2023;00:1–9.

83. Topriceanu CC, Gong X, Shah M, Shiwani H, Eminson K, Atilola GO, Jephcote C, Adams K, Blangiardo M, Moon JC, Hughes AD, Gulliver J, Rowlands A V., Chaturvedi N, O’Regan DP, Hansell AL, Captur G. Higher Aircraft Noise Exposure Is Linked to Worse Heart Structure and Function by Cardiovascular MRI. J Am Coll Cardiol 2025;85:454–469.

84. Raisi-Estabragh Z, Szabo L, Schuermans A, Salih AM, Chin CWL, Vágó H, Altmann A, Ng FS, Garg P, Pavanello S, Marwick TH, Petersen SE. Noninvasive Techniques for Tracking Biological Aging of the Cardiovascular System: JACC Family Series. JACC Cardiovasc Imaging 2024;17:533–551.

85. Salih A, Nichols T, Szabo L, Petersen SE, Raisi-Estabragh Z. Conceptual Overview of Biological Age Estimation. Aging Dis 2023;14:583.

86. Raisi-Estabragh Z, Salih A, Gkontra P, Atehortúa A, Radeva P, Boscolo Galazzo I, Menegaz G, Harvey NC, Lekadir K, Petersen SE. Estimation of biological heart age using cardiovascular magnetic resonance radiomics. Sci Rep 2022;12:12805.

87. Shah M, de A. Inácio MH, Lu C, Schiratti P-R, Zheng SL, Clement A, de Marvao A, Bai W, King AP, Ware JS, Wilkins MR, Mielke J, Elci E, Kryukov I, McGurk KA, Bender C, Freitag DF, O’Regan DP. Environmental and genetic predictors of human cardiovascular ageing. Nat Commun 2023;14:4941.

88. Salih AM, Pujadas ER, Campello VM, McCracken C, Harvey NC, Neubauer S, Lekadir K, Nichols TE, Petersen SE, Raisi-Estabragh Z. Image-Based Biological Heart Age Estimation Reveals Differential Aging Patterns Across Cardiac Chambers. J Magn Reson Imaging 2023;58:1797–1812

89. Mao R, Wang F, Zhong Y, Meng X, Zhang T, Li J. Association of biological age acceleration with cardiac morphology, function, and incident heart failure: insights from UK Biobank participants. Eur Heart J Cardiovasc Imaging 2024;25:1315–1323.

90. Campello VM, Xia T, Liu X, Sanchez P, Martín-Isla C, Petersen SE, Seguí S, Tsaftaris SA, Lekadir K. Cardiac aging synthesis from cross-sectional data with conditional generative adversarial networks. Front Cardiovasc Med 2022;9.

91. Qiao M, McGurk KA, Wang S, Matthews PM, O’Regan DP, Bai W. A personalized time-resolved 3D mesh generative model for unveiling normal heart dynamics. Nat Mach Intell 2025;7:800–811.

92. Raisi-Estabragh Z, Kenawy AAM, Aung N, Cooper J, Munroe PB, Harvey NC, Petersen SE, Khanji MY. Variation in left ventricular cardiac magnetic resonance normal reference ranges: systematic review and meta-analysis. Eur Heart J Cardiovasc Imaging 2021;22:494–504.

93. Parke KS, Brady EM, Alfuhied A, Motiwale RS, Razieh CS, Singh A, Arnold JR, Graham-Brown MPM, Bilak JM, Ayton SL, Dattani A, Yeo JL, McCann GP, Gulsin GS. Ethnic differences in cardiac structure and function assessed by MRI in healthy South Asian and White European people: A UK Biobank Study. J Cardiovasc Magn Reson 2024;26.

94. Ricci F, Aung N, Gallina S, Zemrak F, Fung K, Bisaccia G, Paiva JM, Khanji MY, Mantini C, Palermi S, Lee AM, Piechnik SK, Neubauer S, Petersen SE. Cardiovascular magnetic resonance reference values of mitral and tricuspid annular dimensions: the UK Biobank cohort. J Cardiovasc Magn Reson. 2020;23:5.

95. Raisi-Estabragh Z, Szabo L, McCracken C, Bülow R, Aquaro GD, Andre F, Le TT, Suchá D, Condurache DG, Salih AM, Chadalavada S, Aung N, Lee AM, Harvey NC, Leiner T, Chin CWL, Friedrich MG, Barison A, Dörr M, Petersen SE. Cardiovascular Magnetic Resonance Reference Ranges From the Healthy Hearts Consortium. JACC Cardiovasc Imaging. 2024;

96. Healthy Hearts Consortium. [cited 2024 Dec 7];Available from: https://healthy-hearts.org.uk/

97. Shiwani H, Davies RH, Topriceanu CC, Ditaranto R, Owens A, Raman B, Augusto J, Hughes RK, Torlasco C, Dowsing B, Artico J, Joy G, Miranda I, Witschey W, Rodriguez-Palomares JF, Badia-Molins C, Crotti L, Cortina-Borja M, Chuang ML, Kwong RY, Kramer CM, Manning W, Ho CY, Kellman P, Hughes AD, Biagini E, Mohiddin S, Lopes L, Litt H, Ferrari VA, Captur G, Moon JC, Davies RH, Topriceanu CC, Ditaranto R, Owens A, Raman B, Augusto J, Hughes RK, Torlasco C, Dowsing B, Artico J, Joy G, Miranda I, Witschey W, Rodriguez-Palomares JF, Badia C, Crotti L, Cortina-Borja M, Chuang ML, Kwong RY, Kramer CM, Manning W, Ho CY, Kellman P, Hughes AD, Biagini E, Mohiddin S, Lopes L, Litt H, Ferrari V, Captur G, Moon JC, Bachiller R, Zaha V, Peshock RM, Hobbs HH, Shah A, Rosmini S, Castelletti S, Pierce I, Moschonas K, Webber M, Sivalokanathan S, Malcolmson J, Thompson E, Lovato L, Ponziani A. Demographic-Based Personalized Left Ventricular Hypertrophy Thresholds for Hypertrophic Cardiomyopathy Diagnosis. J Am Coll Cardiol 2025;85:685–695.

98. Hughes RK, Shiwani H, Rosmini S, Augusto JB, Burke L, Jiang Y, Pierce I, Joy G, Castelletti S, Orini M, Kellman P, Xue H, Lopes LR, Mohiddin S, Treibel T, Manisty C, Captur G, Davies R, Moon JC. Improved Diagnostic Criteria for Apical Hypertrophic Cardiomyopathy. JACC Cardiovasc Imaging 2024;17:501–512.

99. Drescher CS, Kelsey MD, Yankey GS, Sun AY, Wang A, Sadeghpour A, Glower DD, Vemulapalli S, Kelsey AM. Imaging Considerations and Clinical Implications of Mitral Annular Disjunction. Circ Cardiovasc Imaging 2022;15:E014243.

100. Zugwitz D, Fung K, Aung N, Rauseo E, McCracken C, Cooper J, El Messaoudi S, Anderson RH, Piechnik SK, Neubauer S, Petersen SE, Nijveldt R. Mitral Annular Disjunction Assessed Using CMR Imaging: Insights From the UK Biobank Population Study. JACC Cardiovasc Imaging 2022;15:1856–1866.

101. McGurk KA, Qiao M, Zheng SL, Sau A, Henry A, Ribeiro ALP, Ribeiro AH, Ng FS, Lumbers RT, Bai W, Ware JS, O’Regan DP. Genetic and phenotypic architecture of human myocardial trabeculation. Nat Cardiovasc Res 2024;3:1503–1515.

102. Petersen SE, Jensen B, Aung N, Friedrich MG, McMahon CJ, Mohiddin SA, Pignatelli RH, Ricci F, Anderson RH, Bluemke DA. Excessive Trabeculation of the Left Ventricle: JACC: Cardiovascular Imaging Expert Panel Paper. JACC Cardiovasc Imaging 2023;16:408.

103. Wang J, Zhang J, Liu W, Pu L, Qi W, Xu Y, Wan K, Gkoutos G V., Han Y, Chen Y. Prognostic Value of Myocardial T1 Mapping for Predicting Adverse Events in Hypertrophic Cardiomyopathy. Circ Cardiovasc Imaging 2025;18:e017174.

104. Raisi-Estabragh Z, McCracken C, Hann E, Condurache D-G, Harvey NC, Munroe PB, Ferreira VM, Neubauer S, Piechnik SK, Petersen SE. Incident Clinical and Mortality Associations of Myocardial Native T1 in the UK Biobank. JACC Cardiovasc Imaging 2023;16:450–460.

105. Nauffal V, Di Achille P, Klarqvist MDR, Cunningham JW, Hill MC, Pirruccello JP, Weng LC, Morrill VN, Choi SH, Khurshid S, Friedman SF, Nekoui M, Roselli C, Ng K, Philippakis AA, Batra P, Ellinor PT, Lubitz SA. Genetics of myocardial interstitial fibrosis in the human heart and association with disease. Nat Genet 2023;55:777.

106. Thanaj M, Mielke J, McGurk KA, Bai W, Savioli N, de Marvao A, Meyer H V., Zeng L, Sohler F, Lumbers RT, Wilkins MR, Ware JS, Bender C, Rueckert D, MacNamara A, Freitag DF, O’Regan DP. Genetic and environmental determinants of diastolic heart function. Nat Cardiovasc Res 2022;1:361–371.

107. Raisi-Estabragh Z, McCracken C, Condurache D, Aung N, Vargas JD, Naderi H, Munroe PB, Neubauer S, Harvey NC, Petersen SE. Left atrial structure and function are associated with cardiovascular outcomes independent of left ventricular measures: a UK Biobank CMR study. Eur Heart J Cardiovasc Imaging 2022:23(9):1191–1200

108. Chadalavada S, Fung K, Rauseo E, Lee AM, Khanji MY, Amir-Khalili A, Paiva J, Naderi H, Banik S, Chirvasa M, Jensen MT, Aung N, Petersen SE. Myocardial Strain Measured by Cardiac Magnetic Resonance Predicts Cardiovascular Morbidity and Death. J Am Coll Cardiol 2024;84:648–659.

109. McCracken C, Szabo L, Abdulelah ZA, Condurache DG, Vago H, Nichols TE, Petersen SE, Neubauer S, Raisi-Estabragh Z. Ventricular volume asymmetry as a novel imaging biomarker for disease discrimination and outcome prediction. Eur Heart J Open 2024;4:oeae059.

110. Aung N, Maciver DH, Zhang H, Chadalavada S, Petersen SE. Global longitudinal active strain energy density (GLASED): a powerful prognostic marker in a community-based cohort. Eur Heart J Cardiovasc Imaging 2024;25:1405–1414.

111. Bonazzola R, Ferrante E, Ravikumar N, Xia Y, Keavney B, Plein S, Syeda-Mahmood T, Frangi AF. Unsupervised ensemble-based phenotyping enhances discoverability of genes related to left-ventricular morphology. Nat Mach Intell 2024;6:291–306.

112. Aung N, Vargas JD, Yang C, Fung K, Sanghvi MM, Piechnik SK, Neubauer S, Manichaikul A, Rotter JI, Taylor KD, Lima JAC, Bluemke DA, Kawut SM, Petersen SE, Munroe PB. Genome-wide association analysis reveals insights into the genetic architecture of right ventricular structure and function. Nat Genet 2022;54:783–791.

113. Benjamins JW, Yeung MW, van de Vegte YJ, Said MA, van der Linden T, Ties D, Juarez-Orozco LE, Verweij N, van der Harst P. Genomic insights in ascending aortic size and distensibility. EBioMedicine 2022;75.

114. Pirruccello JP, Chaffin MD, Chou EL, Fleming SJ, Lin H, Nekoui M, Khurshid S, Friedman SF, Bick AG, Arduini A, Weng LC, Choi SH, Akkad AD, Batra P, Tucker NR, Hall AW, Roselli C, Benjamin EJ, Vellarikkal SK, Gupta RM, Stegmann CM, Juric D, Stone JR, Vasan RS, Ho JE, Hoffmann U, Lubitz SA, Philippakis AA, Lindsay ME, Ellinor PT. Deep learning enables genetic analysis of the human thoracic aorta. Nat Genet 2022;54:40–51.

115. Pirruccello JP, Khurshid S, Lin H, Weng LC, Zamirpour S, Kany S, Raghavan A, Koyama S, Vasan RS, Benjamin EJ, Lindsay ME, Ellinor PT. The AORTA Gene score for detection and risk stratification of ascending aortic dilation. Eur Heart J 2024;45:4318–4332.

116. Pirruccello JP, Rämö JT, Choi SH, Chaffin MD, Kany S, Nekoui M, Chou EL, Jurgens SJ, Friedman SF, Juric D, Stone JR, Batra P, Ng K, Philippakis AA, Lindsay ME, Ellinor PT. The Genetic Determinants of Aortic Distention. J Am Coll Cardiol 2023;81:1320–1335.

117. Francis CM, Futschik ME, Huang J, Bai W, Sargurupremraj M, Teumer A, Breteler MMB, Petretto E, Ho ASR, Amouyel P, Engelter ST, Bülow R, Völker U, Völzke H, Dörr M, Imtiaz MA, Aziz NA, Lohner V, Ware JS, Debette S, Elliott P, Dehghan A, Matthews PM. Genome-wide associations of aortic distensibility suggest causality for aortic aneurysms and brain white matter hyperintensities. Nat Commun 2022;13.

118. Gomes B, Singh A, O’Sullivan JW, Schnurr TM, Goddard PC, Loong S, Amar D, Hughes JW, Kostur M, Haddad F, Salerno M, Foo R, Montgomery SB, Parikh VN, Meder B, Ashley EA. Genetic architecture of cardiac dynamic flow volumes. Nat Genet 2024;56:245–257.

119. Córdova-Palomera A, Tcheandjieu C, Fries JA, Varma P, Chen VS, Fiterau M, Xiao K, Tejeda H, Keavney BD, Cordell HJ, Tanigawa Y, Venkataraman G, Rivas MA, Ré C, Ashley E, Priest JR. Cardiac Imaging of Aortic Valve Area From 34 287 UK Biobank Participants Reveals Novel Genetic Associations and Shared Genetic Comorbidity With Multiple Disease Phenotypes. Circ Genom Precis Med 2020;13:E003014.

120. Yu M, Tcheandjieu C, Georges A, Xiao K, Tejeda H, Dina C, Le Tourneau T, Fiterau M, Judy R, Tsao NL, Amgalan D, Munger CJ, Engreitz JM, Damrauer SM, Bouatia-Naji N, Priest JR. Computational estimates of annular diameter reveal genetic determinants of mitral valve function and disease. JCI Insight 2022;7.

121. Bonazzola R, Ravikumar N, Attar R, Ferrante E, Syeda-Mahmood T, Frangi AF. Image-Derived Phenotype Extraction for Genetic Discovery via Unsupervised Deep Learning in CMR Images. Lecture Notes in Computer Science (including subseries Lecture Notes in Artificial Intelligence and Lecture Notes in Bioinformatics) [Internet]. 2021 [cited 2025 Jul 19];12905 LNCS:699–708. Available from: https://link.springer.com/chapter/10.1007/978-3-030-87240-3_67

122. Burns R, Young WJ, Aung N, Lopes LR, Elliott PM, Syrris P, Barriales-Villa R, Sohrabi C, Petersen SE, Ramírez J, Young A, Munroe PB. Genetic basis of right and left ventricular heart shape. Nat Commun 2024;15:1–17.

123. Aung N, Lopes LR, Van Duijvenboden S, Harper AR, Goel A, Grace C, Ho CY, Weintraub WS, Kramer CM, Neubauer S, Watkins HC, Petersen SE, Munroe PB. Genome-Wide Analysis of Left Ventricular Maximum Wall Thickness in the UK Biobank Cohort Reveals a Shared Genetic Background with Hypertrophic Cardiomyopathy. Circ Genom Precis Med 2023;16:E003716.

124. Yu M, Harper AR, Aguirre M, Pittman M, Tcheandjieu C, Amgalan D, Grace C, Goel A, Farrall M, Xiao K, Engreitz J, Pollard KS, Watkins H, Priest JR. Genetic Determinants of the Interventricular Septum Are Linked to Ventricular Septal Defects and Hypertrophic Cardiomyopathy. Circ Genom Precis Med 2023;16:207–215.

125. Curran L, De Marvao A, Inglese P, McGurk KA, Schiratti PR, Clement A, Zheng SL, Li S, Pua CJ, Shah M, Jafari M, Theotokis P, Buchan RJ, Jurgens SJ, Raphael CE, Baksi AJ, Pantazis A, Halliday BP, Pennell DJ, Bai W, Chin CWL, Tadros R, Bezzina CR, Watkins H, Cook SA, Prasad SK, Ware JS, O’Regan DP. Genotype-Phenotype Taxonomy of Hypertrophic Cardiomyopathy. Circ Genom Precis Med 2023;16:E004200.

126. Zheng SL, Jurgens SJ, McGurk KA, Xu X, Grace C, Theotokis PI, Buchan RJ, Francis C, de Marvao A, Curran L, Bai W, Pua CJ, Tang HC, Jorda P, van Slegtenhorst MA, Verhagen JMA, Harper AR, Ormondroyd E, Chin CWL, Ware JS, de Marvao A, Pantazis A, Baksi J, Halliday BP, Matthews P, Pinto YM, Walsh R, Amin AS, Wilde AAM, Cook SA, Prasad SK, Barton PJR, O’Regan DP, Lumbers RT, Goel A, Tadros R, Michels M, Watkins H, Bezzina CR, Ware JS. Evaluation of polygenic scores for hypertrophic cardiomyopathy in the general population and across clinical settings. Nat Genet 2025;57:563–571.

127. Tadros R, Zheng SL, Grace C, Jordà P, Francis C, West DM, Jurgens SJ, Thomson KL, Harper AR, Ormondroyd E, Xu X, Theotokis PI, Buchan RJ, McGurk KA, Mazzarotto F, Boschi B, Pelo E, Lee M, Noseda M, Varnava A, Vermeer AMC, Walsh R, Amin AS, van Slegtenhorst MA, Roslin NM, Strug LJ, Salvi E, Lanzani C, de Marvao A, Glorioso N, Citterio L, Manunta P, Cusi D, Roberts JD, Tremblay-Gravel M, Giraldeau G, Cadrin-Tourigny J, L’Allier PL, Garceau P, Talajic M, Gagliano Taliun SA, Pinto YM, Rakowski H, Pantazis A, Bai W, Baksi J, Halliday BP, Prasad SK, Barton PJR, O’Regan DP, Cook SA, de Boer RA, Christiaans I, Michels M, Kramer CM, Ho CY, Neubauer S, Woo A, Williamson E, White J, Weintraub W, Weinsaft J, van Rossum A, Sherrid M, Sharma S, Schulz-Menger J, Salerno M, Rimoldi O, Raman B, Plein S, Owens A, Nightingale A, Newby D, Nagueh S, Mongeon FP, Mohiddin S, McCann G, Masri A, Maron M, Mahrholdt H, Mahmod M, Madias C, Larose E, Kwong R, Kolm P, Kim H, Kim B, Kim DY, Jacoby D, Helms A, Heitner SB, Hays A, Geske J, Germans T, Geller N, Gelfand E, Friedrich M, Flett A, et al. Large-scale genome-wide association analyses identify novel genetic loci and mechanisms in hypertrophic cardiomyopathy. Nat Genet 2025;57:530–538.

128. Shah RA, Asatryan B, Sharaf Dabbagh G, Aung N, Khanji MY, Lopes LR, Van Duijvenboden S, Holmes A, Muser D, Landstrom AP, Lee AM, Arora P, Semsarian C, Somers VK, Owens AT, Munroe PB, Petersen SE, Chahal CAA. Frequency, Penetrance, and Variable Expressivity of Dilated Cardiomyopathy-Associated Putative Pathogenic Gene Variants in UK Biobank Participants. Circulation 2022;146:110–124.

129. Jones RE, Hammersley DJ, Zheng S, McGurk KA, de Marvao A, Theotokis PI, Owen R, Tayal U, Rea G, Hatipoglu S, Buchan RJ, Mach L, Curran L, Lota AS, Simard F, Reddy RK, Talukder S, Yoon WY, Vazir A, Pennell DJ, O’Regan DP, Baksi AJ, Halliday BP, Ware JS, Prasad SK. Assessing the association between genetic and phenotypic features of dilated cardiomyopathy and outcome in patients with coronary artery disease. Eur J Heart Fail 2024;26:46–55.

130. Pirruccello JP, Bick A, Wang M, Chaffin M, Friedman S, Yao J, Guo X, Venkatesh BA, Taylor KD, Post WS, Rich S, Lima JAC, Rotter JI, Philippakis A, Lubitz SA, Ellinor PT, Khera A V., Kathiresan S, Aragam KG. Analysis of cardiac magnetic resonance imaging in 36,000 individuals yields genetic insights into dilated cardiomyopathy. Nat Commun 2020;11.

131. Aung N, Nicholls HL, Anwar C, Khanji MY, Rauseo E, Chadalavada S, Petersen SE, Munroe PB, Elliott PM, Lopes LR. Prevalence, Cardiac Phenotype, and Outcomes of Transthyretin Variants in the UK Biobank Population. JAMA Cardiol 2024;9.

132. Alkhodari M, Lapidaire W, Xiong Z, Kart T, Iturria-Medina Y, Hadjileontiadis L, Khandoker A, Lewandowski AJ, Banerjee A, Leeson P. HyperScore: A unified measure to model hypertension progression using multi-modality measurements and semi-supervised learning. Proceedings - 2023 2023 IEEE International Conference on Bioinformatics and Biomedicine, BIBM 2023. 2023;1886–1889.

133. Li Y, Chan E, Puyol-Antón E, Ruijsink B, Cecelja M, King AP, Razavi R, Chowienczyk P. Hemodynamic Determinants of Elevated Blood Pressure and Hypertension in the Middle to Older-Age UK Population: A UK Biobank Imaging Study. Hypertension 2023;80:2473–2484.

134. Hendriks T, Said MA, Janssen LMAA, van der Ende MY, van Veldhuisen DJ, Verweij N, van der Harst P. Effect of Systolic Blood Pressure on Left Ventricular Structure and Function. Hypertension 2019;74:826–832.

135. Kart T, Alkhodari M, Lapidaire W, Banerjee A, Lewandowski AJ, Leeson P. Deep Learning-based Modelling of Complex Hypertensive Multi-Organ Damage with Uncertainty Quantification from Simple Clinical Measures. Proceedings - 2024 IEEE International Conference on Bioinformatics and Biomedicine, BIBM 2024. 2024;1542–1547.

136. Jensen MT, Fung K, Aung N, Sanghvi MM, Chadalavada S, Paiva JM, Khanji MY, de Knegt MC, Lukaschuk E, Lee AM, Barutcu A, Maclean E, Carapella V, Cooper J, Young A, Piechnik SK, Neubauer S, Petersen SE. Changes in Cardiac Morphology and Function in Individuals With Diabetes Mellitus. Circ Cardiovasc Imaging 2019;12:e009476.

137. Bertrand A, Lewis A, Camps J, Grau V, Rodriguez B. Multi-modal characterisation of early-stage, subclinical cardiac deterioration in patients with type 2 diabetes. Cardiovasc Diabetol 2024;23.

138. Brown OI, Drozd M, McGowan H, Giannoudi M, Conning-Rowland M, Gierula J, Straw S, Wheatcroft SB, Bridge K, Roberts LD, Levelt E, Ajjan R, Griffin KJ, Bailey MA, Kearney MT, Cubbon RM. Relationship Among Diabetes, Obesity, and Cardiovascular Disease Phenotypes: A UK Biobank Cohort Study. Diabetes Care 2023;46:1531–1540.

139. Conning-Rowland MS, Giannoudi M, Drozd M, Brown OI, Yuldasheva NY, Cheng CW, Meakin PJ, Straw S, Gierula J, Ajjan RA, Kearney MT, Levelt E, Roberts LD, Griffin KJ, Cubbon RM. The diabetic myocardial transcriptome reveals Erbb3 and Hspa2 as a novel biomarkers of incident heart failure. Cardiovasc Res 2024;120:1898–1906.

140. Petersen SE, Sanghvi MM, Aung N, Cooper JA, Paiva JM, Zemrak F, Fung K, Lukaschuk E, Lee AM, Carapella V, Kim YJ, Piechnik SK, Neubauer S. The impact of cardiovascular risk factors on cardiac structure and function: Insights from the UK Biobank imaging enhancement study. PLoS One. 2017;12:45–52.

141. Aung N, Sanghvi MM, Piechnik SK, Neubauer S, Munroe PB, Petersen SE. The Effect of Blood Lipids on the Left Ventricle. J Am Coll Cardiol. 2020;76:2477–2488.

142. Trudsø LC, Ghouse J, Ahlberg G, Bundgaard H, Olesen MS. Association of PCSK9 Loss-of-Function Variants With Risk of Heart Failure. JAMA Cardiol 2023;8:159–166.

143. Szabo L, McCracken C, Cooper J, Rider OJ, Vago H, Merkely B, Harvey NC, Neubauer S, Petersen SE, Raisi-Estabragh Z. The role of obesity-related cardiovascular remodelling in mediating incident cardiovascular outcomes: a population-based observational study. Eur Heart J Cardiovasc Imaging 2023;24:921–929.

144. van Hout MJP, Dekkers IA, Westenberg JJM, Schalij MJ, Scholte AJHA, Lamb HJ. The impact of visceral and general obesity on vascular and left ventricular function and geometry: a cross-sectional magnetic resonance imaging study of the UK Biobank. Eur Heart J Cardiovasc Imaging 2020;21:273–281.

145. Hartman HS, Kim E, Carbone S, Miles CH, Reilly MP. Sex differences in the relationship between body composition and cardiac structure and function. Eur Heart J Cardiovasc Imaging 2025;26:337–348.

146. Lv Z, Fu Y, Ma Y, Liu C, Yuan M, Gao D. Associations Between Visceral and Liver Fat and Cardiac Structure and Function: A UK Biobank Study. J Clin Endocrinol Metab 2025;110:e1856–e1865.

147. Jamialahmadi O, Tavaglione F, Rawshani A, Ljungman C, Romeo S. Fatty liver disease, heart rate and cardiac remodelling: Evidence from the UK Biobank. Liver Int 2023;43:1247–1255.

148. Roca-Fernandez A, Banerjee R, Thomaides-Brears H, Telford A, Sanyal A, Neubauer S, Nichols TE, Raman B, McCracken C, Petersen SE, Ntusi NA, Cuthbertson DJ, Lai M, Dennis A, Banerjee A. Liver disease is a significant risk factor for cardiovascular outcomes – A UK Biobank study. J Hepatol 2023;79:1085–1095.

149. Ardissino M, McCracken C, Bard A, Antoniades C, Neubauer S, Harvey NC, Petersen SE, Raisi-Estabragh Z. Pericardial adiposity is independently linked to adverse cardiovascular phenotypes: a CMR study of 42 598 UK Biobank participants. Eur Heart J Cardiovasc Imaging 2022;23:1471–1481.

150. Rämö JT, Kany S, Hou CR, Friedman SF, Roselli C, Nauffal V, Koyama S, Karjalainen J, Maddah M, Palotie A, Ellinor PT, Pirruccello JP. Cardiovascular Significance and Genetics of Epicardial and Pericardial Adiposity. JAMA Cardiol 2024;9:418–427.

151. van Meijeren AR, Ties D, de Koning MSLY, van Dijk R, van Blokland I V., Lizana Veloz P, van Woerden G, Vliegenthart R, Pundziute G, Westenbrink DB, van der Harst P. Association of epicardial adipose tissue with different stages of coronary artery disease: A cross-sectional UK Biobank cardiovascular magnetic resonance imaging substudy. IJC Heart and Vasculature 2022;40:101006.

152. Salih A, Ardissino M, Wagen AZ, Bard A, Szabo L, Ryten M, Petersen SE, Altmann A, Raisi-Estabragh Z. Genome-Wide Association Study of Pericardial Fat Area in 28 161 UK Biobank Participants. J Am Heart Assoc 2023;12:e30661.

153. Szabo L, Salih A, Pujadas ER, Bard A, McCracken C, Ardissino M, Antoniades C, Vago H, Maurovich-Horvat P, Merkely B, Neubauer S, Lekadir K, Petersen SE, Raisi-Estabragh Z. Radiomics of pericardial fat: a new frontier in heart failure discrimination and prediction. Eur Radiol 2024;34:4113–4126.

154. Aung N, Sanghvi MM, Zemrak F, Lee AM, Cooper JA, Paiva JM, Thomson RJ, Fung K, Khanji MY, Lukaschuk E, Carapella V, Kim YJ, Munroe PB, Piechnik SK, Neubauer S, Petersen SE. Association Between Ambient Air Pollution and Cardiac Morpho-Functional Phenotypes: Insights From the UK Biobank Population Imaging Study. Circulation 2018;138:2175–2186.

155. Simon J, Fung K, Kolossváry M, Sanghvi MM, Aung N, Paiva JM, Lukaschuk E, Carapella V, Merkely B, Bittencourt MS, Karády J, Lee AM, Piechnik SK, Neubauer S, Maurovich-Horvat P, Petersen SE. Sex-specific associations between alcohol consumption, cardiac morphology and function as assessed by magnetic resonance imaging Insights from the UK Biobank Population Study. Eur Heart J Cardiovasc Imaging 2021;22:1009.

156. Zheng J, Chen H, Yang Q, Zhou Z, Yang C, Huang J, Tu Q, Wu H, Qiu P, Huang W, Shi W, Chen M, Liu H, Shen J, Tang S. Association of coffee consumption and caffeine metabolism with arrhythmias and cardiac morphology: An observational, genetic, and Mendelian randomization study. Heart Rhythm 2025;S1547–5271(24)03631-2

157. Simon J, Fung K, Raisi-Estabragh Z, Aung N, Khanji MY, Kolossváry M, Merkely B, Munroe PB, Harvey NC, Piechnik SK, Neubauer S, Petersen SE, Maurovich-Horvat P. Light to moderate coffee consumption is associated with lower risk of death: a UK Biobank study. Eur J Prev Cardiol 2022;29(6):982–991

158. Evangelou E, Suzuki H, Bai W, Pazoki R, Gao H, Matthews PM, Elliott P. Alcohol consumption in the general population is associated with structural changes in multiple organ systems. Elife 2021;10.

159. Pillinger T, Osimo EF, de Marvao A, Shah M, Francis C, Huang J, D’Ambrosio E, Firth J, Nour MM, McCutcheon RA, Pardiñas AF, Matthews PM, O’Regan DP, Howes OD. Effect of polygenic risk for schizophrenia on cardiac structure and function: a UK Biobank observational study. Lancet Psychiatry 2023;10:98–107.

160. Quiroz JC, Cooper J, McCracken C, Khanji MY, Laranjo L, Aung N, Lee AM, Simon J, Murphy T, Biasiolli L, Piechnik SK, Maurovich-Horvat P, Petersen SE, Raisi-Estabragh Z. The association between adverse childhood experiences and adult cardiac function in the UK Biobank. Eur Heart J Imaging Methods Pract 2024;2.

161. Curta A, Hetterich H, Schinner R, Lee AM, Sommer W, Aung N, Sanghvi MM, Fung K, Lukaschuk E, Cooper JA, Paiva JM, Carapella V, Neubauer S, Piechnik SK, Petersen SE. Subclinical Changes in Cardiac Functional Parameters as Determined by Cardiovascular Magnetic Resonance (CMR) Imaging in Sleep Apnea and Snoring: Findings from UK Biobank. Medicina 2021;57:555.

162. Khanji MY, Karim S, Cooper J, Chahal A, Aung N, Somers VK, Neubauer S, Petersen SE. Impact of Sleep Duration and Chronotype on Cardiac Structure and Function: The UK Biobank Study. Curr Probl Cardiol 2023;48.

163. Thomson RJ, Aung N, Sanghvi MM, Paiva JM, Lee AM, Zemrak F, Fung K, Pfeffer PE, Mackay AJ, McKeever TM, Lukaschuk E, Carapella V, Kim YJ, Bolton CE, Piechnik SK, Neubauer S, Petersen SE. Variation in lung function and alterations in cardiac structure and function—Analysis of the UK Biobank cardiovascular magnetic resonance imaging substudy. PLoS One 2018;13:e0194434.

164. Lv Z, Fu Y, Liu C, Ma Y, Yuan M, Ren J, Gao D. The role of cardiac remodeling associated with renal function in mediating cardiovascular event outcomes. iScience 2024; 27(3):109143.

165. Jamialahmadi O, Tavaglione F, Rawshani A, Ljungman C, Romeo S. Fatty liver disease, heart rate and cardiac remodelling: Evidence from the UK Biobank. Liver Int 2023;43:1247–1255.

166. Condurache DG, D’angelo S, Salih AM, Szabo L, Mccracken C, Mahmood A, Curtis EM, Altmann A, Petersen SE, Harvey NC, Raisi-Estabragh Z. Bone health, cardiovascular disease, and imaging outcomes in UK Biobank: a causal analysis. JBMR Plus 2024;8.

167. Beyer SE, Sanghvi MM, Aung N, Hosking A, Cooper JA, Paiva JM, Lee AM, Fung K, Lukaschuk E, Carapella V, Mittleman MA, Brage S, Piechnik SK, Neubauer S, Petersen SE. Prospective association between handgrip strength and cardiac structure and function in UK adults. PLoS One 2018;13.

168. Raisi-Estabragh Z, Biasiolli L, Cooper J, Aung N, Fung K, Paiva JM, Sanghvi MM, Thomson RJ, Curtis E, Paccou J, Rayner JJ, Werys K, Puchta H, Thomas KE, Lee AM, Piechnik SK, Neubauer S, Munroe PB, Cooper C, Petersen SE, Harvey NC. Poor Bone Quality is Associated With Greater Arterial Stiffness: Insights From the UK Biobank. J Bone Miner Res 2020;36:90–99.

169. Tan CH, Tan JJX. Associations of cardiac function and arterial stiffness with cerebrovascular disease. Int J Cardiol 2024;407:132037.

170. Rauseo E, Salih A, Raisi-Estabragh Z, Aung N, Khanderia N, Slabaugh GG, Marshall CR, Neubauer S, Radeva P, Galazzo IB, Menegaz G, Petersen SE. Ischemic Heart Disease and Vascular Risk Factors Are Associated With Accelerated Brain Aging. JACC Cardiovasc Imaging 2023;16:905–915.

171. Jaggi A, Conole ELS, Raisi-Estabragh Z, Gkontra P, McCracken C, Szabo L, Neubauer S, Petersen SE, Cox SR, Lekadir K. A structural heart-brain axis mediates the association between cardiovascular risk and cognitive function. Imaging Neurosci 2024;2:imag-2-00063.

172. Li X, Yang W, Miao Y, Dove A, Wang J, Du T, Fang Z, Xu W, Zhang Q. Relation of Cognitive Reserve Indicator to Heart Disease and Cardiac Structure and Function: A Large Community-Based Longitudinal Study. J Am Heart Assoc 2024;13.

173. Zhao B, Li T, Fan Z, Yang Y, Shu J, Yang X, Wang X, Luo T, Tang J, Xiong D, Wu Z, Li B, Chen J, Shan Y, Tomlinson C, Zhu Z, Li Y, Stein JL, Zhu H. Heart-brain connections: Phenotypic and genetic insights from magnetic resonance images. Science 2023;380.

174. Amirmoezzi Y, Cropley V, Mansour SL, Seguin C, Zalesky A, Tian YE. Characterizing Brain–Cardiovascular Aging Using Multiorgan Imaging and Machine Learning. J Neurosci 2025;45.

175. van Hout MJP, Dekkers IA, Westenberg JJM, Schalij MJ, Scholte AJHA, Lamb HJ. Associations between left ventricular function, vascular function and measures of cerebral small vessel disease: a cross-sectional magnetic resonance imaging study of the UK Biobank. Eur Radiol 2021;31:5068–5076.

176. Raisi-Estabragh Z, M’Charrak A, McCracken C, Biasiolli L, Ardissino M, Curtis EM, Aung N, Suemoto CK, Mackay C, Suri S, Nichols TE, Harvey NC, Petersen SE, Neubauer S. Associations of cognitive performance with cardiovascular magnetic resonance phenotypes in the UK Biobank. Eur Heart J Cardiovasc Imaging 2021;jeab075.

177. Paik H, Lee J, Jeong CS, Park JS, Lee JH, Rappoport N, Kim Y, Sohn HY, Jo C, Kim J, Cho SB. Identification of a pleiotropic effect of ADIPOQ on cardiac dysfunction and Alzheimer’s disease based on genetic evidence and health care records. Transl Psychiatry 2022;12:1–12.

178. McCracken C, Raisi-Estabragh Z, Veldsman M, Raman B, Dennis A, Husain M, Nichols TE, Petersen SE, Neubauer S. Multi-organ imaging demonstrates the heart-brain-liver axis in UK Biobank participants. Nat Commun 2022;13:1–11.

179. Nauffal V, Klarqvist MDR, Hill MC, Pace DF, Di Achille P, Choi SH, Rämö JT, Pirruccello JP, Singh P, Kany S, Hou C, Ng K, Philippakis AA, Batra P, Lubitz SA, Ellinor PT. Noninvasive assessment of organ-specific and shared pathways in multi-organ fibrosis using T1 mapping. Nature Medicine 2024;30:1749–1760.

